# Repurposed antiviral drugs for COVID-19 –interim WHO SOLIDARITY trial results

**DOI:** 10.1101/2020.10.15.20209817

**Authors:** WHO Solidarity trial consortium, Hongchao Pan, Richard Peto, Quarraisha Abdool Karim, Marissa Alejandria, Ana Maria Henao-Restrepo, César Hernández García, Marie-Paule Kieny, Reza Malekzadeh, Srinivas Murthy, Marie-Pierre Preziosi, Srinath Reddy, Mirta Roses Periago, Vasee Sathiyamoorthy, John-Arne Røttingen, Soumya Swaminathan, as the members of the Writing Committee, assume responsibility for the content and integrity of this article

## Abstract

**BACKGROUND:** WHO expert groups recommended mortality trials in hospitalized COVID-19 of four re-purposed antiviral drugs.

**METHODS:** Study drugs were Remdesivir, Hydroxychloroquine, Lopinavir (fixed-dose combination with Ritonavir) and Interferon-β1a (mainly subcutaneous; initially with Lopinavir, later not). COVID-19 inpatients were randomized equally between whichever study drugs were locally available and open control (up to 5 options: 4 active and local standard-of-care). The intent-to-treat primary analyses are of in-hospital mortality in the 4 pairwise comparisons of each study drug vs its controls (concurrently allocated the same management without that drug, despite availability). Kaplan-Meier 28-day risks are unstratified; log-rank death rate ratios (RRs) are stratified for age and ventilation at entry.

**RESULTS:** In 405 hospitals in 30 countries 11,266 adults were randomized, with 2750 allocated Remdesivir, 954 Hydroxychloroquine, 1411 Lopinavir, 651 Interferon plus Lopinavir, 1412 only Interferon, and 4088 no study drug. Compliance was 94-96% midway through treatment, with 2-6% crossover. 1253 deaths were reported (at median day 8, IQR 4-14). Kaplan-Meier 28-day mortality was 12% (39% if already ventilated at randomization, 10% otherwise). Death rate ratios (with 95% CIs and numbers dead/randomized, each drug vs its control) were: Remdesivir RR=0.95 (0.81-1.11, p=0.50; 301/2743 active vs 303/2708 control), Hydroxychloroquine RR=1.19 (0.89-1.59, p=0.23; 104/947 vs 84/906), Lopinavir RR=1.00 (0.79-1.25, p=0.97; 148/1399 vs 146/1372) and Interferon RR=1.16 (0.96-1.39, p=0.11; 243/2050 vs 216/2050). No study drug definitely reduced mortality (in unventilated patients or any other subgroup of entry characteristics), initiation of ventilation or hospitalisation duration.

**CONCLUSIONS:** These Remdesivir, Hydroxychloroquine, Lopinavir and Interferon regimens appeared to have little or no effect on hospitalized COVID-19, as indicated by overall mortality, initiation of ventilation and duration of hospital stay. The mortality findings contain most of the randomized evidence on Remdesivir and Interferon, and are consistent with meta-analyses of mortality in all major trials. (Funding: WHO. Registration: ISRCTN83971151, NCT04315948)

## INTRODUCTION

A WHO COVID-19 research forum in February 2020 recommended evaluation of treatments in large randomized trials,^1^ and other WHO expert groups identified 4 re-purposed anti-viral drugs that might have at least a moderate effect on mortality: Remdesivir, Hydroxychloroquine, Lopinavir, and Interferon-β1a.^2^ In March 2020, WHO began a large, simple, multi-country, open-label randomized trial among hospital inpatients of the effects of these 4 drugs on in-hospital mortality. The trial was adaptive; unpromising drugs could be dropped and others added. Hydroxychloroquine and Lopinavir were eventually dropped, but others, such as monoclonal antibodies, will be added. We report interim mortality results for the original 4 drugs.

## METHODS

The protocol^3^ was designed to involve hundreds of potentially over-stressed hospitals in dozens of countries. Hence, no form-filling was required, and trial procedures were minimal but rigorous. Online randomization of consented patients (via a cloud-based GCP-compliant clinical data management system) took just a few minutes, as did online reporting of death in hospital or discharge alive (plus brief details of respiratory support in hospital and use of study drugs and certain non-study drugs). No other reporting was required unless doctors suspected an unexpected serious adverse reaction (SUSAR). National and global monitors resolved queries and checked progress and data completeness. Eligible patients were age ≥18 years, hospitalized with a diagnosis of COVID-19, not known to have received any study drug, without anticipated transfer elsewhere within 72 hours, and, in the physician’s view, with no contra-indication to any study drug. Participants were randomized in equal proportions between control and whichever other study drugs were locally available (up to 5 options: these drugs, and local standard-of-care). Placebos were not used. Study drugs were Remdesivir, Hydroxychloroquine, Lopinavir-Ritonavir and Interferon (given with Lopinavir, until July 4). Hydroxychloroquine and Lopinavir were discontinued for futility on June 18 and July 4, 2020, respectively; Interferon is ceasing on October 16.

Daily doses were those already used for other diseases, but to maximize any efficacy without undue cardiac risk Hydroxychloroquine dosage was based on that for amoebic liver abscess, rather than the lower dosage for malaria.^4^ (Hydroxychloroquine slightly prolongs QT, and unduly high or rapid dosage might cause arrhythmias or hypotension.) All treatments were stopped at discharge; otherwise, regimens were:

**Remdesivir (intravenous):** Day 0, 200mg; days 1-9, 100mg.

**Hydroxychloroquine (oral):** Hour 0, four tablets; Hour 6, four tablets; Hour 12, begin two tablets twice daily for 10 days. Each tablet contained 200mg Hydroxychloroquine sulphate (155mg base/tablet; a little-used alternative involved 155mg chloroquine base/tablet).

**Lopinavir (oral):** Two tablets twice daily for 14 days. Each tablet contained 200mg Lopinavir (plus 50mg Ritonavir, to slow hepatic clearance of Lopinavir). Other formulations were not provided, so ventilated patients received no study Lopinavir while unable to swallow.

**Interferon (mainly subcutaneous):** Three doses over six days of 44µg subcutaneous Interferon-ß1a; where intravenous interferon was available, patients on high-flow oxygen, ventilators or ECMO were instead to be given 10µg intravenously once daily for six days.

### ENDPOINTS

The protocol-specified primary objective was to assess effects on in-hospital mortality (ie, mortality during the original episode of hospitalization; follow-up ceased at discharge) not only in all patients but also in those with moderate COVID and in those with severe COVID (subsequently defined as ventilated when randomized).

The protocol-specified secondary outcomes were initiation of ventilation and hospitalization duration. Although no placebos were used, appropriate analyses of these non-fatal outcomes can still be reliably informative. The CATCO add-on study in Canada and the Discovery add-on study in Europe (mostly France) recorded additional outcomes that will be reported elsewhere.

### SAMPLE SIZE

The protocol stated “The larger the number entered the more accurate the results will be, but numbers entered will depend on how the epidemic develops… it may be possible to enter several thousand hospitalised patients with relatively mild disease and a few thousand with severe disease, but realistic, appropriate sample sizes could not be estimated at the start of the trial.” The Executive Group, blind to any findings, decided the timing of release of interim results.

### STATISTICAL ANALYSES

The four main sets of analyses involve the evenly randomized pairwise comparisons of each study drug vs its controls. The controls for those randomly allocated one particular drug were those patients who could by chance have been randomly allocated that drug (at that moment, in that hospital), but instead got allocated standard of care. If, for a particular study entrant, more than one study drug was available, allocation to standard of care would put that patient into the control group for each of them. Hence, there is partial overlap between the four control groups. Each comparison between a study drug and its controls, however, is evenly randomized (50/50) and unbiased, as both groups are affected equally by any differences between countries or hospitals and by any time trends in patient characteristics or standard of care.

All analyses relate mortality to allocated treatment (ie, they are intent-to-treat analyses). The overall mortality analyses were of all randomised patients (drug vs its control), and the only protocol-specified subgroup analyses are those considering separately patients with moderate and with severe COVID (ie, already ventilated; the type of ventilation was not recorded at study entry.)

Unstratified Kaplan-Meier methods plot 28-day risk. Death rate ratios (RRs) and p-values are from log-rank analyses, stratified for 3×2=6 strata of age and ventilation at entry. If the stratified log-rank Observed minus Expected number of deaths is O-E with variance V, logeRR is calculated as (O-E)/V with variance 1/V and a Normal distribution.^8^ The few currently uncertain death times were taken as day 7. Analyses censored patients with outcome not yet reported at day 0, and censored the few inter-hospital transfers at transfer. They did not censor patients discharged alive, as analyses were of mortality during the initial hospitalisation. Forest plots (with 95% CIs only for overall results, otherwise 99% CIs) and chi-squared statistics (sum of [O-E]^2^/V, with no p-value given) help interpret any apparent heterogeneity of treatment RRs between subgroups. Analyses used SASv9.4 and Rv4.02.

The Discussion includes meta-analyses of the major trial results, based on the inverse-variance-weighted average of b=logeRR from each stratum of each trial, using odds ratios where hazard or death rate ratios were unavailable. (This weighted average is derived from the sums of [O-E] and of V over strata.^8^) In general, the more deaths in a stratum the larger V is and, correspondingly, the smaller is the variance of logeRR, so the more weight that stratum gets. The variance attributed to the result in each stratum and to the overall weighted average reflects only the play of chance at randomization. Homogeneity of different RRs is not needed for this weighted average to be informative.

### OVERSIGHT AND FUNDING

The trial is registered (ISRCTN83971151, NCT04315948), with protocol approved by local and WHO ethics committees. Study conduct accorded with Helsinki Declaration and Good Clinical Practice principles, and national trial regulations. Consent forms were signed and retained by patients, but noted for records. Consent was generally prospective but could (where locally approved) be retrospective. The only exclusions were patients without clear consent to follow-up. All other randomized patients were included (“intent-to-treat analyses”). WHO is global co-sponsor and governments national co-sponsors, with trial governance by the International Steering Committee’s Executive Group (EG). External statistical analyses for the independent Data and Safety Monitoring Committee (DSMC) were unseen by the EG or WHO, with two exceptions. After outside evidence of Hydroxychloroquine and Lopinavir futility, the EG requested unblinded analyses of them. Second, after deciding blindly to report all interim results, the EG revised this manuscript, drafted by the WHO trial team and external statisticians.

Participating countries covered almost all local costs and WHO covered all other study costs, receiving no extra funding. Collaborators, committee members, data analysts and data management systems charged no costs, and drugs were donated. No donor unduly (see end-material) influenced analyses, manuscript preparation, or submission. The Writing Group vouch for protocol fidelity and data accuracy and completeness.

## RESULTS

From March 22 to October 4, 2020, 11,330 patients were entered from 405 hospitals in 30 countries in all 6 WHO regions. Of these, 64 (0.6%) had no, or uncertain, consent to follow-up, leaving 11,266 for intent-to-treat analyses: 2750 allocated Remdesivir, 954 Hydroxychloroquine, 1411 only Lopinavir-ritonavir, 2063 Interferon, and 4088 no study drug (Figure 1; reporting is 97% complete for those entered >1 month earlier, and 99.7% complete for those entered >3 months earlier). All 3 patients with COVID refuted are included, and survived.

**Figure 1.**
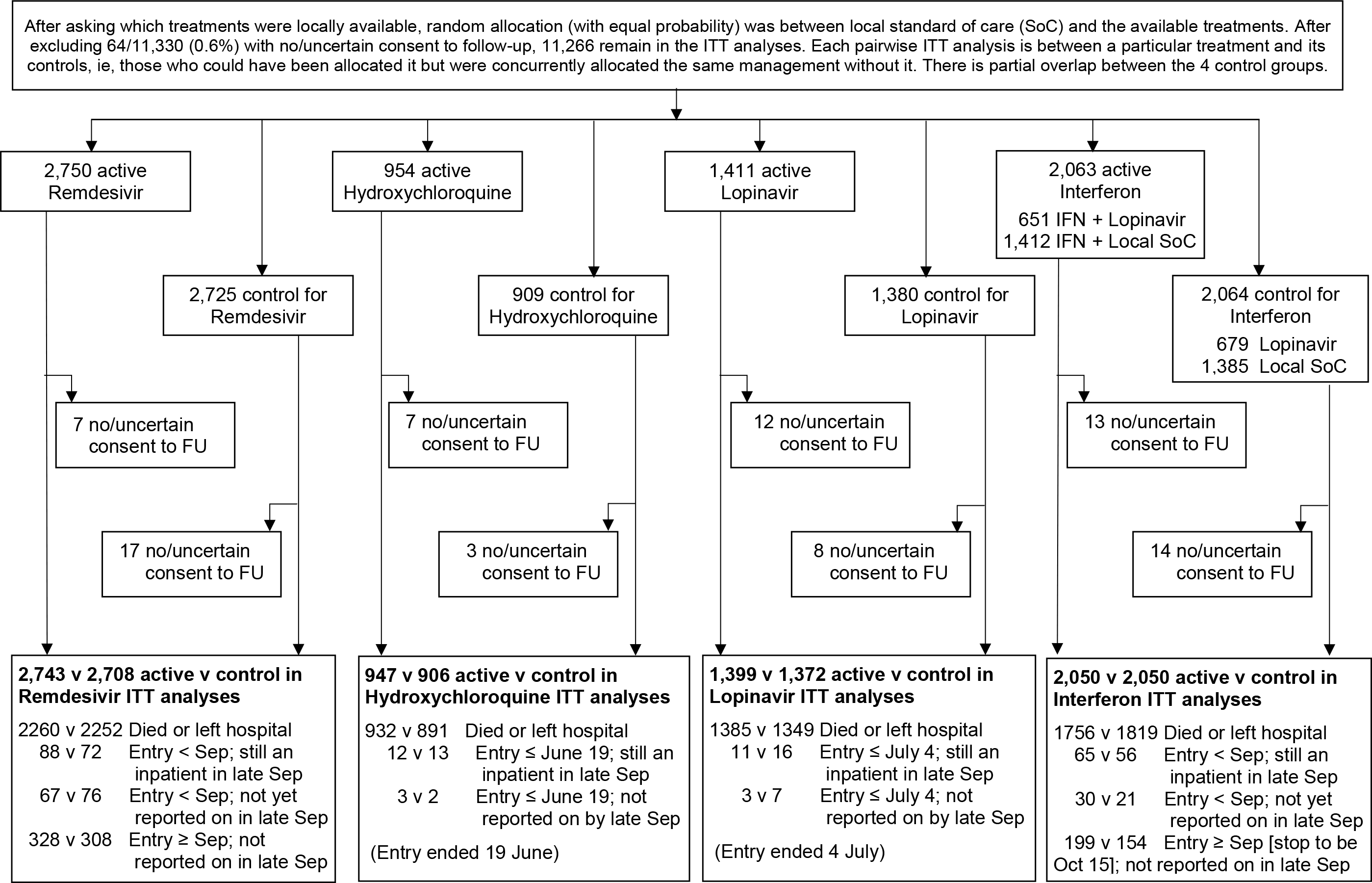
WHO Solidarity Trial – information to October 4, 2020 on entry, follow-up (FU) and intent-to-treat (ITT) analyses

Table 1 shows patient characteristics: 9120 (81%) age <70 years, 6985 (62%) male, 2768 (25%) with diabetes, 916 (8%) already ventilated, and 7002 (62%) randomized on days 0-1. For each drug, patient characteristics were well balanced by the unstratified 50/50 randomization between it and its controls. Deaths were at median day 8 (IQR 4-14) and discharges at median day 8 (IQR 5-13). With 1253 deaths, the Kaplan-Meier estimate of 28-day mortality was 11.8%. This risk depended on several factors, particularly age (20% if ≥70 years, 6% if <50 years) and ventilation (39% if ventilated, otherwise 10%).

**Table 1.**
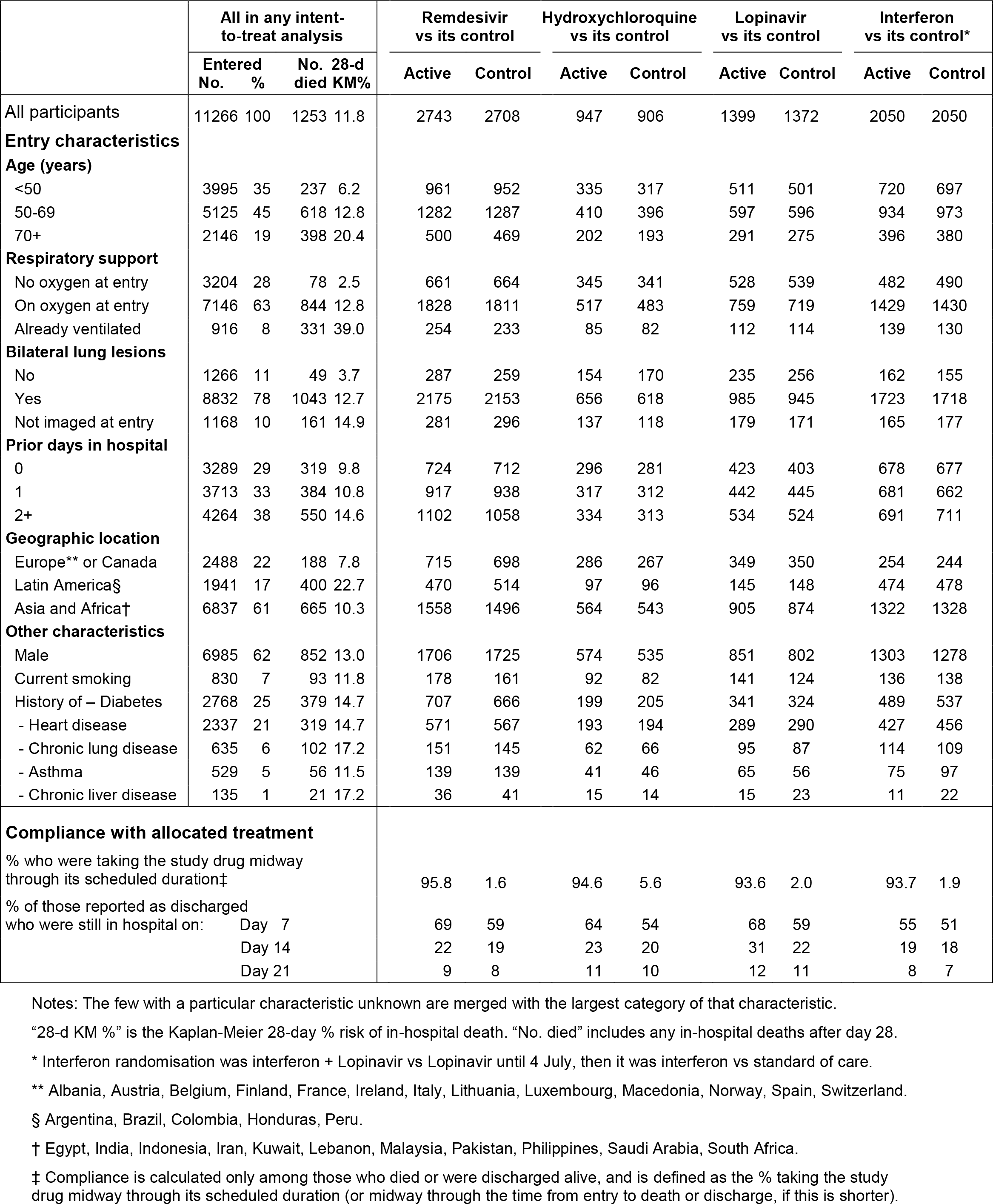
Entry characteristics by random allocation, and compliance with that allocation. Excludes 64 without clear consent to follow-up. Comparisons are of each study drug vs concurrent allocation to the same treatment without it. As the control groups overlap, the total number (11,266) is less than the sum of the numbers in the pairwise comparisons.

Table 1 also describes compliance. For Remdesivir the scheduled treatment period was 10 days (or to prior death or discharge). Of those allocated Remdesivir, 98.5% began treatment. Midway through this period 96% were still taking it (as against only 2% of the Remdesivir controls). Likewise, for other drugs compliance midway was high (94-95%) and crossover low (2-6%). Study treatments ceased on schedule. Absolute treatment vs control differences in use of corticosteroids and other non-study treatments were small (Table S2).

For each pairwise drug comparison, Figure 2 gives unstratified Kaplan-Meier analyses of 28-day in-hospital mortality (listing below the x-axis numbers at risk and dying in each week, and numbers dying after day 28), along with death rate ratios (RRs) stratified for age and ventilation. Figure 3 gives RRs in subgroups of age stratified by ventilation and of ventilation stratified by age, and overall RRs stratified by both.

**Figure 2.**
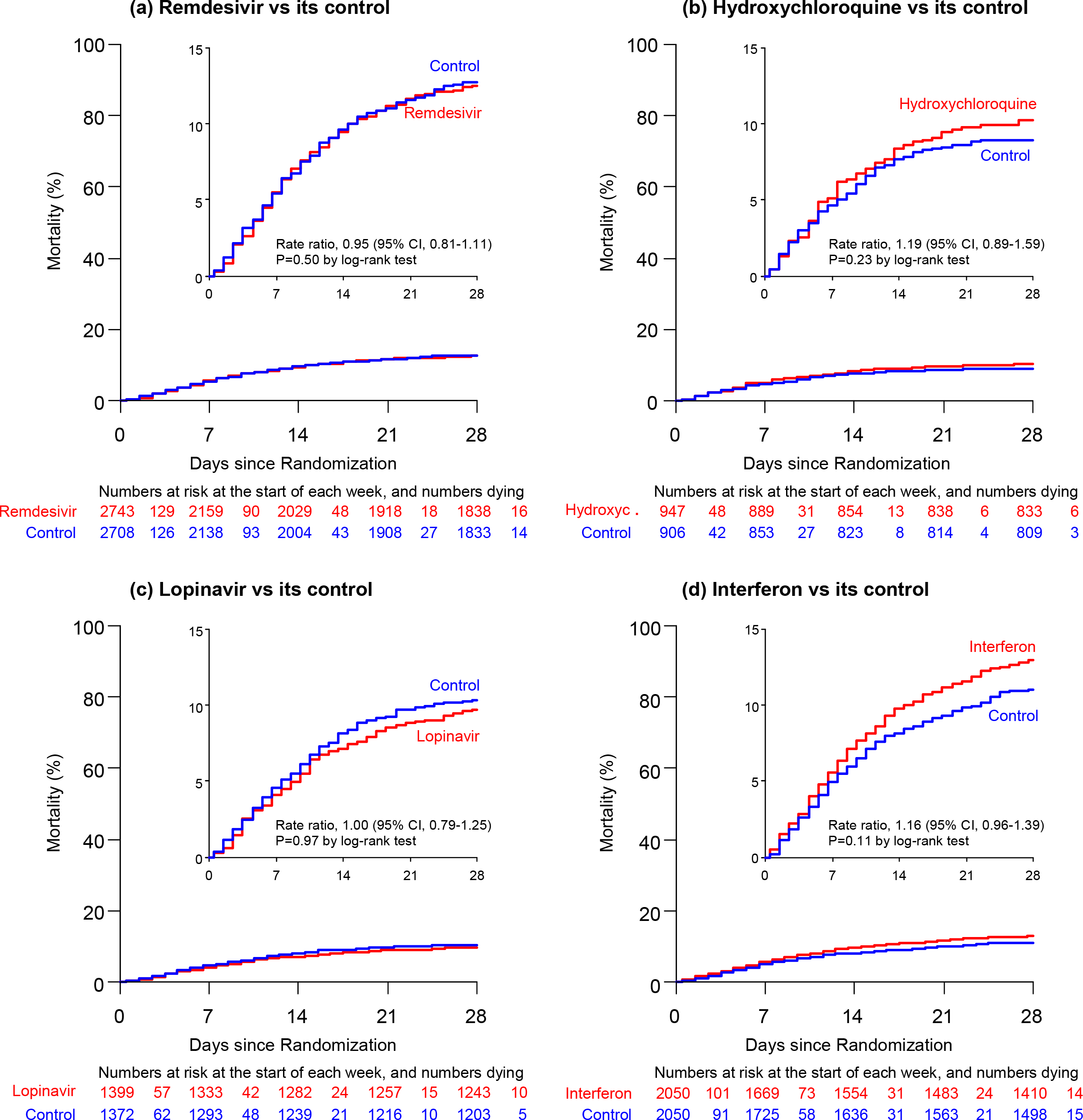
Effects of (a) Remdesivir, (b) Hydroxychloroquine, (c) Lopinavir, and (d) Interferon on 28-day mortality. Kaplan-Meier graphs of in-hospital mortality. The inset shows the same data on an expanded y-axis.

**Figure 3.**
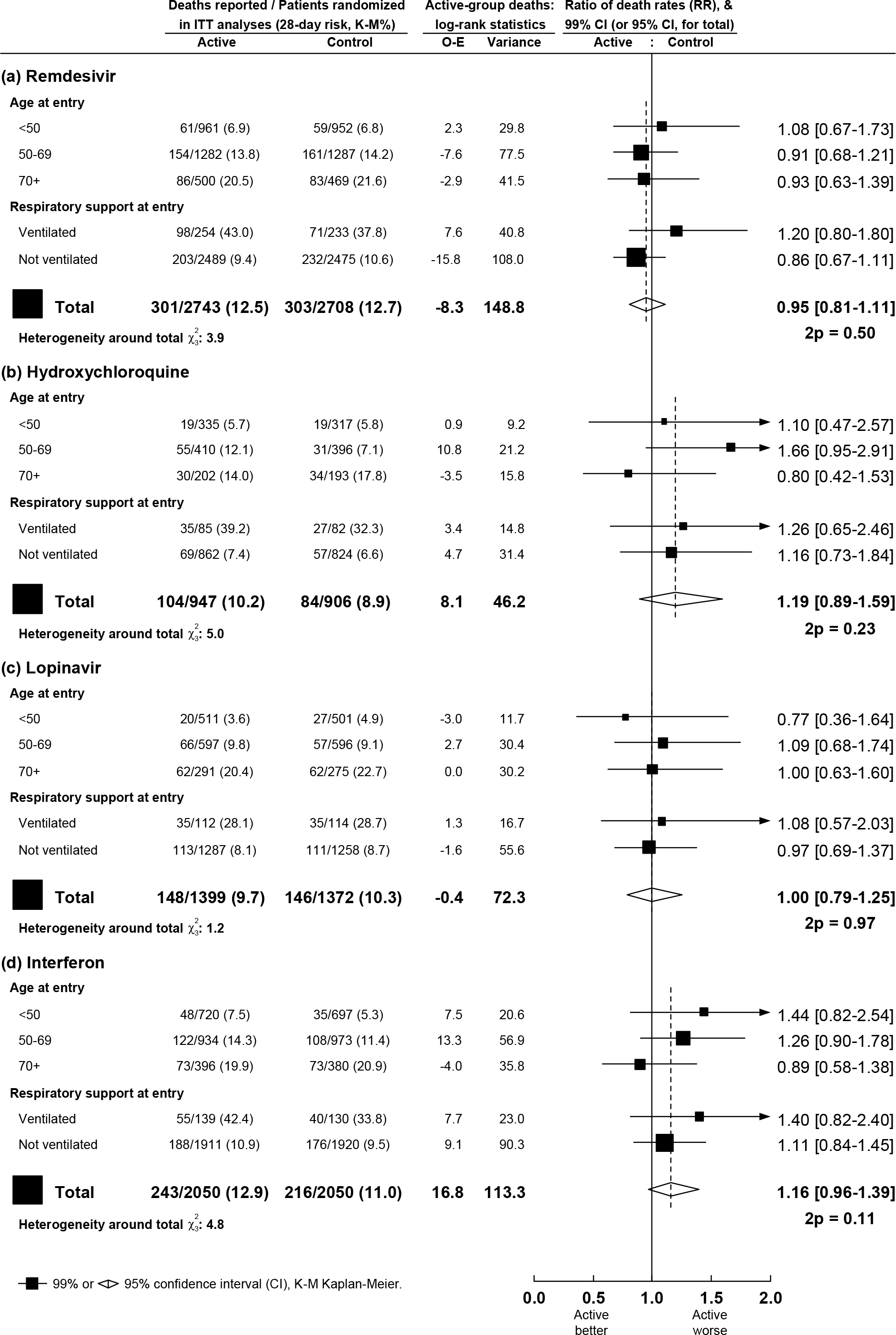
Rate ratios of any death, stratified by age and respiratory support at entry, (a) Remdesivir, (b) Hydroxychloroquine, (c) Lopinavir, (d) Interferon, each vs its control

Taking Figures 2 and 3 together, no study drug had any definite effect on mortality, either overall (each p>0.10) or in any subgroup defined by age or ventilation at entry (or other entry characteristics, or geographic region, or corticosteroid use: Figures S6-S9). Death rate ratios (with 95% CIs, and drug vs control numbers of deaths thus far reported) were: Remdesivir RR=0.95 (0.81-1.11, p=0.50; 301/2743 vs 303/2708), Hydroxychloroquine RR=1.19 (0.89-1.59, p=0.23; 104/947 vs 84/906), Lopinavir RR=1.00 (0.79-1.25, p=0.97; 148/1399 vs 146/1372) and Interferon RR=1.16 (0.96-1.39, p=0.11; 243/2050 vs 216/2050). Unstratified comparisons yielded similarly null findings (Figure 2), as did analyses excluding corticosteroid users and multivariate sensitivity analyses estimating simultaneously the effects of all 4 study drugs (Table S3).

If ventilation prevents oral administration of Lopinavir or other study drugs then this could reduce any effects on mortality of allocation to those drugs, but the pre-planned analyses of mortality in patients not already ventilated at entry also indicated no definite protective effect of any study drug (Figure 3).

The pre-planned study outcomes were death, ventilation and time to discharge. No study drug appreciably reduced initiation of ventilation in those not already ventilated. The numbers, study drug vs control, with ventilation initiated after randomization were: Remdesivir 295v284, Hydroxychloroquine 75v66, Lopinavir 124v119, Interferon 209v210 (Table S1).

In this open-label trial, patients who would be considered fit for discharge might be kept in somewhat longer just because they were being given a study drug, but this difficulty can be circumvented. Each of the 3 study treatments scheduled to last >7 days *increased* the percentages remaining in hospital at day 7. If one of these 3 drugs had accelerated recovery then the sizes of these increases should have differed, but they did not: the increases were strikingly similar. The proportions still hospitalized at day 7, study drug vs control, were Remdesivir 69%v59%, Hydroxychloroquine 64%v54%, Lopinavir 68%v59% (Table 1). The medically informative result is the lack of any material difference between these 3 increases.

Supplementary analyses by treatment allocation (Tables S2-S3, Figures S2-S9) tabulate co-medication (finding only small absolute differences), provide a multi-variable Cox regression fitting all 4 treatment effects simultaneously (yielding mortality RRs like those in Figure 3), subdivide 28-day mortality graphs (like those in Figure 2) by ventilation at entry, and give subgroup analyses of mortality RRs by many patient characteristics and by corticosteroid use (identifying no noteworthy subgroup-specific or geographic variation).

All active treatment ended within ≤14 days, and the numbers of deaths during this 14-day period with any cardiac cause mentioned on the electronic death record was Remdesivir 7v8, Hydroxychloroquine 4v2, Lopinavir 6v3, and Interferon 6v8 (Figure S11). Although many COVID deaths involved multi-organ failure, no study drug death was attributed to renal or hepatic disease.

## DISCUSSION

The main outcomes of mortality, initiation of ventilation and hospitalization duration were not clearly reduced by any study drug. The mortality findings cannot have been appreciably biased by the open-label design without placebos, or by variation in patient characteristics or local care. The effects on ventilation initiation are unlikely to have been materially biased, and although allocation to 10 days of medication can delay discharge while medication is being given, the striking similarity of this delay with 3 different daily medications is evidence that none had a pharmacological effect that appreciably reduced time to recovery. Although ACTT-1, with placebo control, reported Remdesivir moderately reduced time to recovery, in the present study there were no material effects on ventilation initiation or time to discharge.

The chief aim was to help determine whether any of 4 re-purposed antivirals could at least moderately affect in-hospital mortality, and whether any effects differed between moderate and severe disease. The results should be considered in the context of all the evidence on mortality from properly randomized trials, but for Remdesivir and for Interferon this study provides more than three-quarters of that evidence.

There are 4 trials of Remdesivir vs the same management without it: Solidarity (604 deaths in about 5000 randomized), ACTT-1 (136 deaths in about 1000) and two smaller trials (41 deaths).^5-7^ Figure 4 gives mortality results from each trial, subdivided by initial respiratory support. (These like-vs-like comparisons allow for the proportion already on high-flow oxygen or ventilation at entry into ACTT-1 having been, by chance, somewhat lower with Remdesivir than with placebo.) Combining data appropriately from all 4 trials,^8^ the Remdesivir vs control death rate ratio (RR) is 0.91 (95% CI 0.79-1.05).

**Figure 4.**
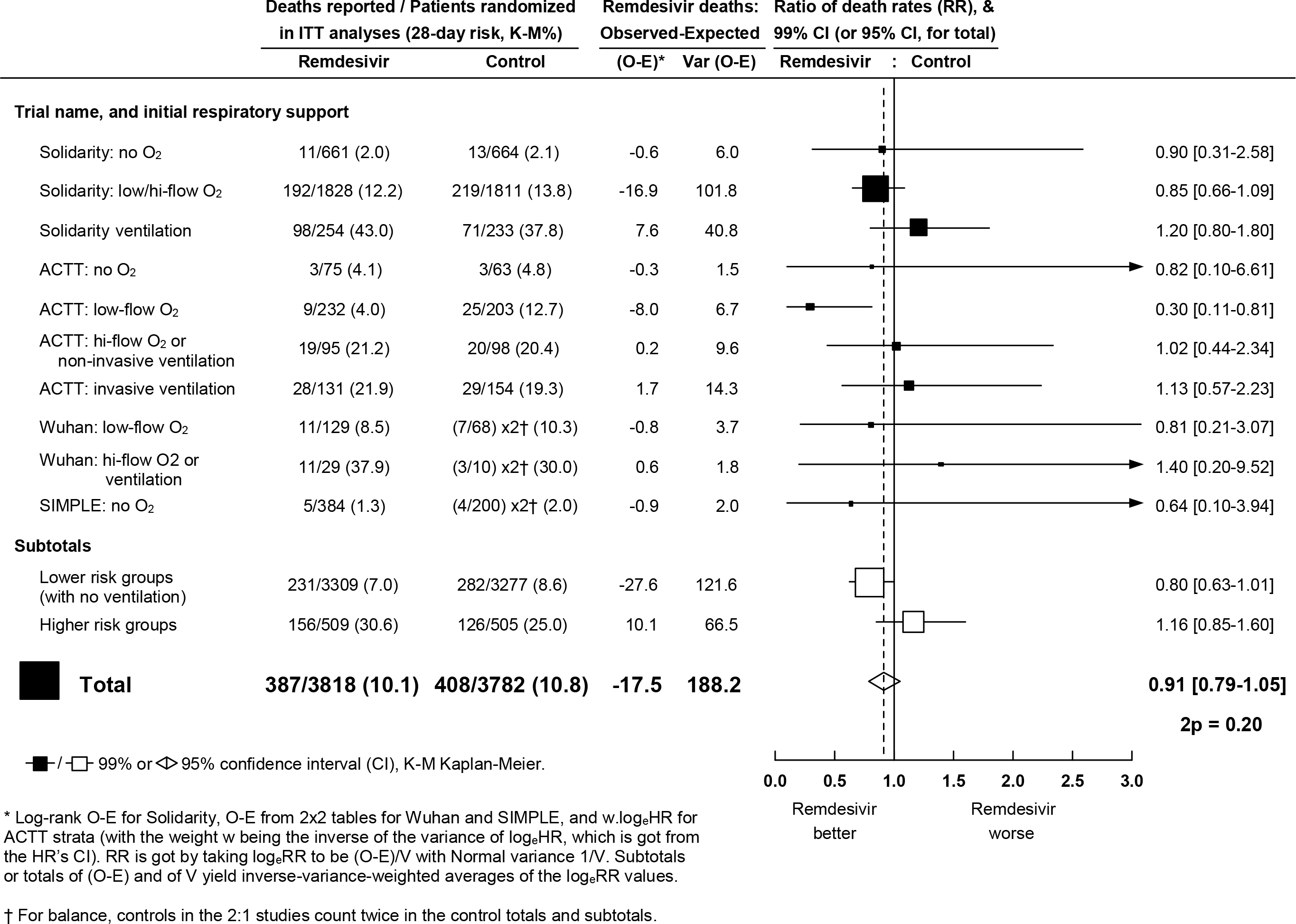
Remdesivir vs control – Meta-analysis of mortality in trials of random allocation of hospitalised COVID-19 patients to Remdesivir or the same treatment without it

Interpretation should chiefly reflect not the p-value (p=0.21) or point estimate (RR=0.91) but the confidence interval (0.79-1.05), which shows the range of death rate ratios comfortably compatible with the weighted average of the findings from all trials. This absolutely excludes the suggestion that Remdesivir can prevent a substantial fraction of all deaths. The confidence interval is comfortably compatible with prevention of a small fraction of all deaths, but is also comfortably compatible with prevention of no deaths (which would be consistent with the apparent lack of any reduction by Remdesivir in the initiation of ventilation or the duration of hospitalization in Solidarity).

The statistical uncertainties are much greater if attention is restricted to particular subgroups or time periods.^9^ If Remdesivir has no effect on mortality then chance could still produce somewhat favourable findings in a subgroup of the results for all trials, with more striking findings in a selected subgroup of a particular trial (as in the one subgroup of ACTT-1 where the death rate ratio appeared to be 0.30: Figure 4). Although both ACTT-1 and Solidarity envisaged the possibility of different degrees of benefit in lower- and higher-risk patients, the particular lower-risk/higher-risk subdivision of the ACTT-1 findings in Figure 4 was unplanned. (The ACTT-1 protocol specified separate analyses of those not requiring *any* oxygen, with only 3/75 vs 3/63 deaths in ACTT-1, 11/661 vs 13/664 in Solidarity, and 5/384 vs 4/200 in SIMPLE; overall RR=0.82, but with wide confidence interval 0.43-1.55.) Thus, although the all-trials subtotals in Figure 4 suggest some benefit in low-risk patients and some hazard in high-risk inpatients (with the absolute benefit in low-risk appearing somewhat smaller than the absolute hazard in high-risk), neither subtotal should be considered in isolation from the other subtotal, or from the CI for the total.

For Hydroxychloroquine and Lopinavir, Solidarity found no definite evidence of benefit or of hazard in any subgroup. The only other substantial trial of these two drugs is Recovery,^10,11^ which for these drugs is larger than Solidarity. Combination of log-rank analyses from these two relatively large trials (by the meta-analysis methods used in Figure 4) consolidate the findings of both.

For Hydroxychloroquine, the joint mortality RR (combining 2 trials) was 1.11, 95% CI 0.99-1.24, with no apparent benefit whether ventilated or not. This CI excludes any material benefit from this Hydroxychloroquine regimen in hospitalized COVID. It is compatible with some hazard, but does not demonstrate hazard. Despite concerns that the loading dose could be temporarily cardiotoxic, in neither trial was there any excess mortality during the first few days, when blood levels were highest. Neither trial recorded dosage/kg, obesity, or cardiac parameters, and cardiac deaths were too few to be reliably informative. A recent meta-analysis identified 27 small randomized Hydroxychloroquine trials (total 167 deaths, RR=1.00, 0.71-1.42);^12^ combining all 29 trials, RR=1.10, 0.99-1.22, again excluding any material benefit.

For Lopinavir (always co-administered with Ritonavir), the joint mortality RR (combining Solidarity, Recovery and the only informative smaller trial^13^) was 1.02, 95% CI 0.91-1.14. Although Lopinavir tablets could not be swallowed by ventilated patients, there was no apparent benefit in analyses restricted to those not already being ventilated at entry. This CI indicates no material effect on mortality, and excludes a 10% proportional reduction. An add-on study within Solidarity, Discovery, recorded many clinical parameters, identifying an unexpected increase in creatinine (perhaps because blood levels are higher than in similarly-dosed HIV patients^14,15^), but Solidarity and Recovery recorded no renal or hepatic deaths with Lopinavir.

For Interferon-β1a no large mortality trials have been reported. Based on about 4000 patients, the mortality RR in Solidarity was 1.16, 0.96-1.39; p=0.11 (or 1.12, 0.83-1.51, without Lopinavir co-administration: Figure S9). This does not demonstrate hazard, but the lower confidence limit does exclude a moderate mortality reduction in these circumstances. About half the interferon-allocated patients (and half their controls) received corticosteroids,^16^ but the interferon vs control mortality RR seemed unaffected by corticosteroids. Most interferon was subcutaneous, and subcutaneous and intravenous interferon have different pharmacokinetics,^17^ but the clinical relevance of this is unclear. Randomization to Interferon is ceasing in Solidarity on October 16, but other evidence will emerge: a report that nebulized Interferon-β1a might be highly effective involved only about 100 COVID patients (NCT04385095), but the ongoing placebo-controlled ACTT-3 trial of subcutaneous Interferon-β1a aims to involve 1000 (NCT04492475).

For each of these 4 repurposed non-specific antivirals, several thousand patients have now been randomized in various trials. The unpromising overall findings from the regimens tested suffice to refute early hopes, based on smaller or non-randomized studies, that any will substantially reduce inpatient mortality, initiation of ventilation or hospitalisation duration. Narrower confidence intervals would be helpful (particularly for Remdesivir), but the main need is for better treatments. Solidarity is still recruiting about 2000 patients per month, and efficient factorial designs will allow it to assess further treatments, such as immune-modulators and specific anti-SARS-Cov-2 monoclonal antibodies.

The chief acknowledgement is to the thousands of patients and their families who participated in this trial, and the hundreds of medical staff who randomized and cared for the patients. The Ministries of Health of the participating Member States and their national institutions provided critical implementation support. The views expressed are those of the Writing Group, not necessarily those of WHO. NJ White et al^4^ provided unpublished Hydroxychloroquine pharmacokinetic data, the Recovery trial^10,11^ shared log-rank statistics, the ACTT-1 trial^5^ shared subgroup hazard ratios, and Bin Cao shared Wuhan trial^6^ details.

MS preparation, revision and submission was controlled by the WHO trial team and the writing committee. There were no funders for the main Solidarity trial, but the Discovery add-on study received EU Horizon 2020 research and innovation program grant 101015736. Participating countries covered almost all local costs and WHO covered all other study costs, receiving no extra funding. Collaborators, committee members, data analysts and data management systems charged no costs, and drugs were donated. Castor EDC donated and managed their cloud-based clinical data capture and management system, blind to study findings. Anonymized data handling and analysis was by the Universities of Berne, Bristol and Oxford. Remdesivir was donated by Gilead Sciences, Hydroxychloroquine by Mylan, Lopinavir by Abbvie, Cipla and Mylan, and Interferon β1a by Merck KGaA (subcutaneous) and Faron (intravenous).

## Data Availability

These are interim results. Once the final database is locked, data sharing requests will be considered.

