## Supplementary appendix for "Repurposed antiviral drugs for COVID-19 –interim WHO SOLIDARITY trial results"

This page will be deleted before publication, and replaced by the Journal’s cover page

### Contents list for supplementary online material

**Page**


**3** Membership: Writing Committee; Data and Safety Monitoring Committee

**4** Membership: International Steering Committee and its Executive Group

**5** National investigators and researchers

**12** Data managers, statistical analysts and WHO trial co-ordination team

**12** Other collaborators in participating countries

**14** Acknowledgements

**15** Supp. Table S1. Treatment allocation vs respiratory support in hospital in patients not ventilated at entry and eventually discharged alive

**16** Supp. Table S2. Use of corticosteroids and other non-study drugs

**17** Supp. Table S3. Multivariate analysis simultaneously estimating all 4 treatment effects

**18** Supp. Fig S1. Effects on 28-day in-hospital mortality of (a) Remdesivir, (b) Hydroxychloroquine, (c) Lopinavir, (d) Interferon

**19** Supp. Fig S2. Effects of Remdesivir on the 28-day risk of death from any cause, subdivided by Respiratory support at entry

**20** Supp. Fig S3. Effects of Hydroxychloroquine on the 28-day risk of death from any cause, subdivided by Respiratory support at entry

**21** Supp. Fig S4. Effects of Lopinavir on the 28-day risk of death from any cause, subdivided by Respiratory support at entry

**22** Supp. Fig S5. Effects of Interferon on the 28-day probability of death from any cause, subdivided by Respiratory support at entry

**23** Supp. Fig S6. RRs of any death, Remdesivir vs Control, by entry characteristics

**24** Supp. Fig S7. RRs of any death, Hydroxychloroquine vs Control, by entry characteristics

**25** Supp. Fig S8. RRs of any death, Lopinavir vs Control, by entry characteristics

**26** Supp. Fig S9. RRs of any death, Interferon vs Control, by entry characteristics

**27** Supp. Fig S10. RRs for the composite outcome of death or initiation of ventilation

**28** Supp. Fig S11. Treatment allocation vs cardiac death

#### Writing Committee

Hongchao Pan, Ph.D., Richard Peto, F.R.S., Quarraisha Abdool Karim, Ph.D., Marissa Alejandria M.D., M.Sc., Ana Maria Henao-Restrepo, M.D., M.Sc., César Hernández García M.D., Ph.D., Marie Paule Kieny Ph.D., Reza Malekzadeh M.D., Srinivas Murthy M.D. C.M., Marie-Pierre Preziosi M.D., Ph.D., K. Srinath Reddy M.D., D.M., Mirta Roses Periago M.D., MPH, Vasee Sathiyamoorthy B.M.B.Ch., Ph.D., John-Arne Røttingen M.D., Ph.D., Soumya Swaminathan M.D.

Nuffield Department of Population Health, University of Oxford, Oxford, United Kingdom (H.P. and R.P.), Centre for the AIDS Programme of Research In South Africa (CAPRISA), Durban, South Africa (Q.A.K.), National Institutes of Health, University of the Philippines, Manila, Philippines (M.M.A.), Agency of Medicine and Medical Devices, Madrid, Spain (C.H.G), Institut National de la Santé Et de la Recherche Médicale (INSERM), Paris, France (M.P.K.), Digestive Disease Research Institute, Teheran University of Medical Sciences, Tehran, Iran (R.M.), University of British Columbia, Vancouver, Canada (S.M), Public Health Foundation of India, New Delhi, India (K.S.R.), National Academy of Sciences of Buenos Aires, Buenos Aires, Argentina (M.R.P.), Research Council of Norway, Oslo, Norway (J-A.R.), World Health Organization, Geneva, Switzerland (A-M.H-R., M-P.P., V.S.M., S.S.).

#### Global Data and Safety Monitoring Committee

Aldo Maggioni (chair), Abdel Babiker, Deborah Cook, Arjen Dondorp, Gagandeep Kang

#### International Steering Committee

**National Principal Investigators; †National Coordinators; ‡Members of the Executive Group.*

**Albania:** *University Hospital Centre Mother Theresa, Tirana* N Como*; *National Agency for Medicines and Medical Devices* N Sinani†. **Argentina:** *Fundación del Centro de Estudios Infectológicos* G Lopardo*; *National Academy of Sciences of Buenos Aires* M Roses Periago†‡. **Austria:** *Through DISCOVERY add-on study.* **Belgium:** *Through DISCOVERY add-on study.* **Brazil:** *Oswaldo Cruz Foundation* EP Nunes*, PPS Reges†. **Canada:** *University of British Columbia* S Murthy*‡; *Public Health Agency of Canada* M Salvadori†. **Colombia:** *National University of Colombia* CA Alvarez- Moreno*; *Ministry of Health* ML Mesa Rubio†.
**Egypt:** *National Hepatology and Tropical Medicine Research Institute* M Hassany*; *Ministry of Health and population* H Zaid†. **Finland:** *Helsinki University Hospital and South Karelian Central Hospital, Lappeenranta* KAO Tikkinen*; *Finnish Institute for Health and Welfare and University of Finland, Helsinki* M Perola†. **France:** *Through DISCOVERY add-on study. Hospices Civils de Lyon, Lyon* F Ader*; *Institut National de la Santé Et de la Recherche Médicale, Paris* MP Kieny†‡. **Honduras:** *National Autonomous University of Honduras* MT Medina*; *Secretaria de Salud de Honduras* N Cerrato†. **India**: *ICMR,National AIDS Research Institute, Pune* S Godbole*†; *Public Health Foundation of India* KS Reddy‡. **Indonesia**: *National Institute of Health Research and Development* I Irmansyah*; *RSUP Persahabatan, Jakarta* MR Rasmin†. **Iran (Islamic Republic of):** *Digestive Disease Research Institute, Teheran University of Medical Sciences, Tehran* R Malekzadeh*†‡. **Ireland**: *HRB Clinical Research Facility, University College Cork* J Eustace*; *Department of Health* T Maguire†. **Italy**: *University of Verona* E Tacconelli*; *Italian Medicines Agency (AIFA)* N Magrini†. **Kuwait:** *Infectious Diseases Hospital* A Alhasawi*; *Ministry of Health* A Al-Bader†. **Lebanon:** *Rafic Hariri University Hospital* P Abi Hanna*; *Ministry of Public Health* R Hamra†. **Lithuania:** *University Hospital Santaros Klinikos, Vilnius* L Jancoriene*, L Griskevicius†. **Luxembourg**: *Through DISCOVERY add-on study.* **Malaysia**: *Penang Hospital* TS Chow*; *Hospital Sungai Buloh, Jalan Hospital* S Kumar†. **North Macedonia**: *University Clinic of Infectious Diseases and Febrile Conditions* M Stevanovikj*; *Ministry of Health* S Manevska†. **Norway**: *Oslo University Hospital* P Aukrust*, A Barratt-Due†; *Research Council of Norway* JA Røttingen‡. **Pakistan:** *Shaukat Khanum Memorial Cancer Hospital and Research Centre* M Hassan*†. **Peru:** *Universidad Peruana Cayetano Heredia* PJ García*, E Gotuzzo†. **Philippines:** *National Institutes of Health, University of the Philippines, Manila* MM Alejandria*†‡. **Saudi Arabia**: *Ministry for Preventive Health* AO Athari Alotaibi*, A Asiri†. **South Africa:** *University of the Witwatersrand* J Nel*; *Wits Reproductive Health and HIV Institute* H Rees†; *Centre for the AIDS Programme of Research In South Africa* Q Abdool Karim‡. **Spain:** *Hospital Clinico San Carlos, UCM, IdISSC, Madrid* A Portoles*; *Agency of Medicine and Medical Devices* C Hernández-Garcia†‡. **Switzerland:** *Lausanne University Hospital* O Manuel*†.

#### Executive Group of the International Steering Committee

John-Arne Røttingen (chair), Quarraisha Abdool Karim, Marissa Alejandria, César Hernández García, Marie Paule Kieny, Reza Malekzadeh, Srinivas Murthy, K. Srinath Reddy, Mirta Roses Periago, Soumya Swaminathan, Richard Peto (independent statistician).

#### National investigators and researchers

*This does not include members of the International Steering Committee or its Executive Group.*

**Albania:** *University Hospital Centre Mother Theresa, Tirana* NGJ Gjermeni, E Meta.

**Argentina**: *Health Ministry* JB Balbuena, JM Castelli, A Mykietiuk, C Vizzotti; *Hospital de Infecciosas Francisco J Muñiz, Buenos Aires* V Chediack, E Cunto, L de Vedia, C Domínguez, J Fernández, N Lista, A Rodríguez; *Hospital General de Agudos José Ramos Mejía, Buenos Aires* S Caimi, C Delgado, M Losso, F Masciottra, V Pachioli, J Toibaro; *Hospital General de Agudos Juan A Fernández, Buenos Aires* J Barletta, J Carrillo, N D'Amico, L Hermida, M Jaume, C Luna, M Padilla, J Patroso, L Perez Blanco, J Presas, MJ Rolon, AL Sisto, S Themines; *Hospital Julio C Perrando, Resistencia, Chaco* V Arce, P Arribillaga, RA Ferreyra, ML Lescano, F Tito, L Verón; *Hospital Mariano y Luciano de la Vega, Moreno* A Chalco, J Farina, M Provenzano; *Hospital Nacional Profesor Alejandro Posadas, Palomar* I Alonso, RA Alzola, M Benedetti, D Di Pilla, P Díaz Aguiar, C Giudiche, M Golikow, M Jacobo, D Laplume, F Loiacono, A López, L Olleros, C Pallavicini, F Riveros, G Torales; *Hospital Prof. Bernado Houssay, Vicente López* M Altamirano, L Barcelona, V Berdiñas, C Fogar, A Martin; *Hospital Provincial Dr José María Cullen, Santa Fé* RA Avila, J Burgui, N Carrizo, M Filippi, M Gomez, V Reichert; *Hospital Rawson, Córdoba* M Alvarez, AC Cazaux, M Díaz, HM Hurtado, LR Lorena, ML Marianelli, L Orellano, C Salvay, M Simonetta.

**Austria**: *Through DISCOVERY add-on study. Paracelsus Medical University Salzburg, SCRI-CCCIT and AGMT* A Egle, R Greil; *Medizinische Universität Innsbruck, Innsbruck* M Joannidis.

**Belgium:** *Through DISCOVERY add-on study. CHR de la Citadelle, Liège* A Altdorfer, V Fraipont; *Cliniques Universitaires de Saint Luc, Bruxelles* L Belkhir; *Hôpital Erasme, Bruxelles* M Hites.

**Brazil:***Fundação Universidade de Pernambuco* DB Miranda Filho, P Monteiro; *Hospital Couto Maia* VPS Almeida, CX Nunes; *Hospital das Clínicas da Universidade Federal de Minas Gerais* H Duani; *Hospital das Clínicas da Universidade Federal do Paraná* GL Breda, SM Raboni; *Hospital Estadual de Sumaré* AJS Colussi, MC Ramos, LF Ruffing; *Hospital Federal do Estado do Rio de Janeiro* EC João; *Hospital Regional de Mato Grosso do Sul* JHC Croda; *Hospital Regional de São José* GA Pinto; *Hospital São José de Doenças Infecciosas* EAG Arruda; *Hospital Sírio Libanês* MFDB Corradi; *Hospital Universitário Clementino Fraga Filho* ES Machado, FCQ Mello; *Instituto de Infectologia Emilio Ribas* LC Pereira Junior, TNL Souza, ALC Toscano; *Oswaldo Cruz Foundation* VGV Santos.

**Canada:** *Centre Hospitalier de l'Universite de Montreal* FM Carrier, M Durand; *Centre Hospitalier Universitaire de Sherbrooke* F Lamontagne; *CHU de Quebec-Universite Laval* D Bellemare, E Cloutier, O Costerousse, TV Tran, A Turgeon; *Grey Nuns Community Hospital* H Hoang; *Hopital Montfort* N Chagnon; *IUCPQ* F Lellouche; *Lions Gate Hospital* J Douglas; *Markham Stouffville Hospital*, E Fera; *McGill University, Montreal* MP Cheng, C Costiniuk, L Harrison, K Khwaja, M Klein, N Kronfli, TC Lee, J Papenburg, M Semret; *McMaster university* E Duan; *Memorial University of Newfoundland* T Azher; *North York General Hospital, Toronto* A Geagea; *Ottawa Hospital* S English; *Queen's University* S Perez-Patrigeon; *Queensway Carleton Hospital* M Rushton; *Royal Alexandra Hospital* A Singh; *Royal Victoria Regional Health Centre* G DiDiodato; *Sinai Health System, Toronto* M Fralick; *St Paul's Hospital* N Press; *Sunnybrook Hospital* R Fowler, A Rishu; *Trillium Health Partners* C Graham; *University Health Network, Toronto* I Bogocj; *University of Alberta* N Lee, C O'Neil; *University of British Columbia* D Ovakim; *University of Calgary, Calgary* J Conly, CD Fell, R Lim, R Somayaji, A Tremblay, E Vakil; *University of Manitoba* Y Keynan, R Zarychanski; *Vancouver General Hospital* A Mah; *Western University* S Parvathy, M Silverman; *William Osler Health System* A Binnie, S Borgia.

**Colombia:** *Clínica Colsanitas, Sede Clínica Iberomérica* I Zuluaga; *Clínica Colsanitas, Sede Clínica Reina Sofia* J Chacón, D Garzón, F Guevara; *Clínica Colsanitas, Sede Clinica Santa Maria del Lago* JS Bravo; *Clínica Colsanitas, Sede Clínica Sebastian de Belalcazar* JM Oñate; *Clínica Colsanitas, Sede Clínica Universitaria Colombia* S Lozano-González; JA Rojas-Murrugarra, CS Saavedra; *Fundación Cardio Infantil, Iinstituto del Corazón* EV Váquiro, F Varón-Vega; *Fundación Hospital Universidad del Norte* H Macareno; *Fundación Santa Fe de Bogotá* M Caicedo; *Fundación Valle de Lili* F Rosso; *Hospital Universitario San Ignacio, Pontificia Universidad Javeriana* SL Valderrama.

**Egypt:** *Ain Shams University* G Elassal; *AL Azhar University* S Zaki; *Assuit University* S Hassany, E Moustafa; *Cairo University* A Abdalmohsen, A Abdelbary, N Asem, H Masoud; *Ministry of Health and Population,* W Amin, M Elshesheny, M Fathy, N Fathy, N Fayed, A Hammam, M Solyman Kabyl, M Mohamed, A Mohamed Gouda, S Okasha, A Rafik, A Sedky, S Tarek, A Tharwat; *National Hepatology and Tropical Medicine Research Institute* A Abdel Baki; *National Liver Institute* W Abdel-Razek; *National Research Center* E Kamal.

**Finland:** *Helsinki University Hospital, Helsinki* M Myllärniemi, J Paajanen, A Renner; *Tampere University Hospital, Tampere* J Rutanen, MU Sinisalo.

**France**: ***Through DISCOVERY add-on study.*** *Amiens University, Amiens* C Andrejak, JP Lanoix, Y Zerbib; *ANRS, Paris* A Diallo, N Mercier; *Centre hospitalier Andrée Rosemon, Cayenne, Guyane* F Djossou; *Centre Hospitalier Annecy Genevois, Annecy* D Bougon, V Tolsma; *Centre Hospitalier Universitaire de Besançon, Besançon* K Bouiller, JC Navellou; *Centre Hospitalier Universitaire de Nantes, Nantes* B Gaborit, F Raffi, J Reignier; *Centre Hospitalier Universitaire Dijon-Bourgogne, Dijon* P Andreu, L Piroth, JP Quenot; *Centre Hospitalier Universitaire Grenoble Alpes, Grenoble* O Epaulard, N Terzi; *Centre Régional Universitaire de Nancy, Vandoeuvre Lés Nancy* F Goehringer, A Kimmoun; *Centre Régional Universitaire de Nice, Nice* J Courjeon, J Dellamonica, S Leroy, CH Marquette; *Centre Régional Universitaire de Rennes, Rennes* F Laine, B Laviolle, *Georges Pompidou European Hospital, Paris* D Lebeaux, *Groupe Hospitalier de la région Mulhouse Sud Alsace, Mulhouse* O Hinschberger, Y Mootien; *Groupe hospitalier La Pitié-Salpêtrière, Paris* J Mayaux, V Pourcher; *Groupe Hospitalier Paris Saint Joseph, Paris* C Bruel, B Pilmis; *Henri-Mondor Hospital, Créteil* S Gallien, A Mekontso Dessap; *Hôpital Bichat, Paris* T Alfaiate, D Belhadi, L Bouadma, C Burdet, A Dechanet, A Dupont, S Laribi, FX Lescure, F Mentre, N Peiffer-Smadja, G Peyvatin, MC Tellier, JF Timsit, Y Yazdanpanah; *Hôpital Cochin, Paris* S Kerneis, M Lachatre, O Launay; *Hôpital de Bicêtre, Le Kremlin Bicêtre* S Figueiredo, S Jauréguiberry; *Hôpital Delafontaine, Saint Denis* J Aboab, F Crockett, N Sayre; *Hôpital d'instruction des armées Bégin, Saint Mandé* C Dubost); *Hôpital Marie Lannelongue, Le Plessis Robinson* J Le Pavec, F Stefan; *Hôpital Saint-Antoine, Paris* K Lacombe; *Hôpital Saint-Louis, Paris* JM Molina, M Noret; *Hôpital Tenon, Paris* G Pialoux; *Hospices Civils de Lyon, Lyon* JC Richard, J Textoris, F Wallet; *Institut National de la Santé Et de la Recherche Médicale, Paris* C Delmas, J Saillard; *Lapeyronie University Hospital, Montpellier* K Klouche; *Lille University Hospital, Lille* K Faure, E Faure, J Poissy; *Metz-Thionville hospital, Ars-Laquenexy* R Gaci, C Robert; *Montpellier University Hospital, Montpellier* V Le Moing, A Makinson; *Pontchaillou University Hospital, Rennes* F Benezit; *Reims University Hospital,* B Mourvillier ; *Sorbonne Université, Paris* D Costagliola; *Strasbourg University Hospital, Strasbourg* R Clere-Jehl, F Danion , F Meziani, V Poindron; *Toulouse University Hospital, Toulouse* F Bounes, G Martin-Blondel; *Tourcoing Hospital, Tourcoing* V Jean-Michel, E Senneville; *Tours University Hospital, Tours* D Garot; *University Hospital Centre of Bordeaux, Bordeaux,* C Cazanave, D Gruson, D Malvy; *University Hospital of Martinique, Fort-de-France* C Chabartier; *University Hospital of Saint-Etienne, Saint-Etienne* E Botelho-Nevers, A Gagneux-Brunon, G Thiery; *University Hospital, Rennes* C Fougerou.

**Honduras:** *Hospital Atlantida, la Ceiba* AA Fiallos; *Hospital Leonardo Martinez, San Pedro Sula* L Erazo; *Hospital Militar, Tegucigalpa* R Figueroa; *Hospital San Felipe, Tegucigalpa* JJ Flores, L Melendez; *Instituto Cardiopulmonar, Tegucigalpa* C Aguilar, W Moncada.

**India**: *AIIMS, Bhopal* S Atal, R Joshi, S Khadanga, A Ray, S Saigal, S Sharma; *AIIMS, Jodhpur* A Avinash, P Bhardwaj, P Bhatia, J Charan, N Chauhan, N Dutt, M Garg, V Nag, B Shadrach; *AIIMS, New Delhi* R Aggarwal, DK Baidya, , R Guleria, CA Kayina, A Mittal, N Nischal, M Soneja, K Soni, S Maitra, A Trikha, N Wig; *AIIMS, Rishikesh* G Chikara, P Gupta, R Kant, V Krishnan, B Mohan, P Panda; *Apollo Hospitals, Greams Lane, Chennai* N Ramakrishnan, BK Tirupakuzhi Vijayaraghavan, R Venkatasubramanian; *Apollo Speciality Hospitals, Vanagaram, Chennai* R Ebenezer, S Krishnamoorthy, D Suresh Kumar; *Army Institute of Cardio Thoracic Sciences, Pune* G Bhati, V Marwah, D Peter, TVSVGK Tilak; *B. J. Government Medical College & Sassoon General Hospital, Pune* R Borse, B Daswani, S Divhare, D Ogale, S Sangale, M Tambe, R Waghmare; *B.J. Medical College& New Civil Hospital, Ahmedabad* C Desai, D Raval, K Upadhyay; *Bharati hospital, Pune* N Agrawal, S Iyer, K Reddy, S Rege, J Shah; *BYL Nair Hospital, Mumbai* R Bhadade, R de Souza, M Harde; *Chirayu Medical College & Hospital, Bhopal* A Goenka, A Mangalgiri, M Maurya, R Parate, K Singh, A Tiwari, R Verma; *Christian Medical College, Vellore* OC Abraham, A Balachandran, TD Sudarsanam; *Gandhi Hospital, Hyderabad* (V Aedula, T C Bingi, V Jamalapuram, H Kalakuntla, A K Maurya, K Nagmani, K Padma Malini, M Rajarao, KT Rao, R Sudarsi, M D Suleman; *GMERS Medical College & Hospital, Gotri, Vadodara* K Mehta, P Patel, C Rathod; *Government Medical College and New Civil Hospital, Surat* C Acharya, K Bhatt, M Chaudhari, V Chaudhary, B Divakar, A Gamit, S Gamit, B Kantharia, A Kavishvar, M Momin, C Patel, V Patel, S Patel, H Patel, A Vasava, M Verma; *Government Medical College, Nagpur* S Khandare, D Chand, M Kalikar, S Mitra, U Narlawar; *Government. Siddhartha Medical College, Vijayawada* B Bhargavi, G Chakradhararao, D Durgaprasad, K Seshaiah; *ICMR- National AIDS Research Institute, Pune* S Chidrawar, A Kadam, S Kalme, S Kamble, M Mamulwar, S Panda, S Sane; *Indian Council of Medical Research, New Delhi* B Bhargava, R Gangakhedkar, N Gupta; *Madras Medical College & Rajiv Gandhi Government General Hospital, Chennai* T Banu, V Damodaran, L Narasimhan, G Natarajan, V Rajendran, KM Sudha, S Sudharshini, E Therani; *Rajan Omandurar Medical College & Hospital, Chennai* R Jayanthi, J Komathi, KP Manimaran, T Ramesh Kumar, A Revathi; *Pandit Deendayal Upadhyay Government Medical College, Rajkot* M Bhapal, S Misra, A Singh, A Trivedi); *PD Hinduja National Hospital and Medical Research Centre, Mumbai* U Agrawal, Z Udwadia; *RCSM GMC CPRH, Kolhapur* A Paritekar, G Patil, A Waikar; *Sardar Vallabhbhai Patel Institute of Medical Sciences and Research, Ahmedabad* S Malhotra, D Roy; *SMS Medical College & Hospital, Jaipur* A Agrawal, S Bhandari, S Mahavar, R Sharma, S Sharma, A Singh; *Voluntary Health Services-Infectious Diseases Medical Centre, Chennai* N Kumarasamy, P Selvamuthu; *WHO-India, New Delhi* M Ahmad, M Gupta, V Purohit.

**Indonesia**: *National Institute of Health Research and Development* AR Afrilia, D Arlinda, R Avrina, LE Bang, SL Driyah, M Erastuti, T Fajarwati, M Karyana, N Nurhayati, C Opitasari, AA Pradana, Y Risniati, RI Sugiyono, NH Susanto, AK Syarif, A Yulianto; *RS University Airlangga, Surabaya* M Amin; *RS University Udayana Bali* IKA Somia, *RS YARSI, Jakarta* I Kusuma; *RSJ Prof. Dr. Soerojo, Magelang* HA Mahmudji; *RSPAU Dr. Esnawan Antariksa, Jakarta* FE Sari; *RSPI Prof. Dr. Sulianto Saroso, Jakarta* PA Sitompul; *RSUD Dr. Achmad Mochtar, Bukittinggi* D Herman; *RSUD Dr. Moewardi, Solo* H Harsini; *RSUD Dr. Saiful Anwar, Malang* YJ Sugiri; *RSUD Dr. Soetomo, Surabaya* S Soedarsono; *RSUP Dr. Hasan Sadikin, Bandung* Y Hartantri; *RSUP Dr. Kariadi, Semarang* SB Raharjo; *RSUP Dr. M. Djamil, Padang* I Medison; *RSUP Dr. Sardjito, Yogyakarta* BS Riyanto; *RSUP Dr. Wahidin Sudirohusodo, Makassar* I Djaharuddin; *RSUP Fatmawati, Jakarta* AY Djojo; *RSUP H. Adam Malik, Medan* A Rahmaini; *RSUP Persahabatan, Jakarta* F Isbaniah; *RSUP Prof. Dr. R. D Kandou Manado* A Nugroho; *RSUP Sanglah, Bali* GK Sajinadiyasa; *RSUPN Dr. Cipto Mangunkusumo, Jakarta* CW Pitoyo.

**Iran (Islamic Republic of):** *Ahvaz Jundishapur University of Medical Sciences, Ahvaz* F Amini, S Moogahi, M Varnasseri, MJ Yadyad, F Yousefi; *Alborz University of Medical Sciences, Karaj* Z Siami, A Soleimani; *Arak University of Medical Sciences, Arak* A Kamali, B Mahmoodiyeh, H Sarmadian, D Shojaei, S Soltanmohammad; *Babol University of Medical Sciences, Babol* M Bayani, S Ebrahimpour, M Javanian, M Sadeghi Haddad Zavareh, M Shokri; *Golestan University of Medical Sciences, Gorgan* B Khodabakhshi, A Norouzi, S Tavassoli; *Guilan University of Medical Sciences, Rasht* F Joukar, L Mahfoozi, F Mansour-Ghanaei, A Pourkazemi; *Isfahan University of Medical Sciences, Isfahan* A Hakamifard, M Salahi, K Shirani; *Kermanshah University of Medical Sciences, Kermanshah* M Afsharian, A Janbakhsh, F Mansouri, R Miladi, P Mohamadi, Z Mohseni Afshar, B Sayad, M Shirvani, S Vaziri, MH Zamanian; *Mashhad University of Medical Sciences, Mashhad* M Amini, F Barazandeh, S Hafizi Lotfabadi, R Khodashahi, M Mozdourian, SN Saberhosseini, M Saberi, N Saber-Moghaddam, Y Yazdanpanah; *Mazandaran University of Medical Sciences, Sari* F Baba Mahmoodi, F Fallahpoor Golmaee; National Institute for Medical Research Development, Tehran B Mesgarpour; *Qazvin University of Medical Sciences, Qazvin* A Karampour, S Kiani Majd, R Najafipour, H Najari, E Zare Hoseinzade; *Qom University of Medical Sciences, Qom* SY Foroghi Ghomi, MR Ghadir, M Gheitani, SS Hashemi Madani, A Hormati, J Khodadadi; *Saveh University of Medical Sciences, Saveh* A Akhavi Mirab, M Mesri, H Mozaffar; *Shahid Beheshti University of Medical Sciences, Tehran* P Baghaei, F Dastan, P Tabarsi; *Shahid Sadoughi University of Medical Sciences, Yazd* SA Mousavi Anari; *Shiraz University of Medical Sciences, Shiraz* MJ Fallahi, M Moghadami, S Yaghoubi, F Zand; *Tabriz University of Medical Sciences, Tabriz* K Ansarin, H Mikaeili, M Nazemiyeh, A Taghizadieh; *Tehran University of Medical Sciences, Tehran* S Eghtesad, F Ghiasvand, H Hosseini, N Khajavirad, M Mohraz, H Poustchi, A Sadeghi, MA Sahraian, MR Salehi, AR Sima.

**Ireland**: *Beaumont Hospital and Royal College of Surgeons in Ireland* E deBarra; *Mater Misericordiae University Hospital* E Muldoon; *Mercy University Hospital* A Jackson; *St James's Hospital and Trinity College, Dublin* C Bergin; *St Vincent's University Hospital and School of Medicine University College Dublin* C McCarthy; *University Hospital Galway* JG Laffey.

**Italy**: *AOU Citta della Salute e Scienza, Torino* S Corcione, FG De Rosa, S Scabini; *ASST di Monza, Ospedale San Gerardo, Monza* L Bisi, P Bonfanti, G gustinetti, F Iannuzzi; *ASST Fatebenefratelli Sacco* A Capetti, M Galli, S Rusconi; *ASST Santi Paolo e Carlo, Milano* F Bai, A d’Arminion Monforte, E Merlini; *ASST Valtellina e Alto Lario, Ospedale di Sondalo* E Menatti, P Zucchi; *Azienda Ospedaliera Ospedali Riuniti Marche Nord, Pesaro* F Barchiesi, B Canovari; *Azienda Sanitaria Universitaria Friuli Centrale (ASU FC)* D Pecori, C Tascini, P Della Siega, M Merelli; *Azienda Socio Sanitaria Territoriale di Cremona* N Cocco, B Drera, C Fornabaio, A Pan; *Brescia Spedali Civili General Hospital* F Castelli, E Focà, E Roldan; *Fondazione IRCCS Ca' Granda Ospedale Maggiore Policlinico, Milan* A Bandera, A Gori; *Fondazione Policlinico Universitario A. Gemelli IRCCS* R Cauda, A Cingolani, K de Gaetano Donati, S Lamonica; *IRCSS Ospedale Sacro Cuore – Don Calabria, Negrar Di Valpolicella (Verona)* A Agheben, N Riccardi, P Rodari; *Ospedale Cardinal Massaia, Asti* M Degioanni, T Lupia; *Ospedal Maggiore* S Di Bella, D Giacomazzi, R Luzzati; *Ospedale Policlinico San Martino – IRCCS* M Bassetti; *Ospedale SM Goretti, Latina* B Kertusha, M Lichtner, P Zuccalá; *Policlinico di S. Orsola, Bologna* C Campoli, P Viale; *ULSS9 Scaligera, Verona* P Rovere, M Vincenzi; *Unità Operativa Dipartimentale di Malattie Infettive* M Vincenzi; *University of Verona* E Cremonini, P De Nardo, MD Pezzani.

**Kuwait:** *Infectious Diseases Hospital* M Al-Roomi, S Kelly; *Kuwait University* S Al-Sabah.

**Lebanon :** *Centre Hospitalier Universitaire, Notre Dame des Secours* M Matar; *Rafic Hariri University Hospital* M Hassoun, M Saliba.

**Lithuania:** *University Hospital Santaros Klinikos, Vilnius* B Zablockiene.

**Luxembourg**: *Through DISCOVERY add-on study. Centre Hospitalier de Luxembourg* J Reuter, T Staub.

**Malaysia**: *Queen Elizabeth Hospital* HG Lee; *Institute for Clinical Research, National Institute of Health* CK Chew, PP Goh, WY Mak; *Kuala Lumpur Hospital* CL Leong; *Melaka Hospital* NZ Zaidan; *Sarawak General Hospital* HH Chua; *Sultanah Bahiyah Hospital* LL Low; *Sungai Buloh Hospital* YG Mohamed Gani; *Tengku Ampuan Afzan Hospital* D Muhamad; *Tuanku Fauziah Hospital* S Ab Wahab.

**North Macedonia**: *University Clinic of Infectious Diseases and Febrile Condition* IS Demiri, S Marinkovikj, B Petreska, K Spasovska.

**Norway**: *Akershus University Hospital* O Dalgard; *Diakonhjemmet Hospital* L Vinge; *Haraldsplass Deaconess Hospital* BR Kittang; *Haukeland University Hospital* B Blomberg; *Innlandet Hospital, Elverum* CM Ystrøm; *Innlandet Hospital, Lillehammer* R Eiken; *Levanger Hospital* NV Skei; *Lovisenberg Hospital* H Hoel; *Molde Hospital* B Tholin; *Møre og Romsdal Hospital* DAL Hoff; *Oslo University Hospital* AM Dyrhol-Riise, AR Holten, T Kåsine, K Nezvalova-Henriksen, IC Olsen , M Trøseid; *Østfold Hospital* S Aballi; *Sorlandet Hospital, Arendal* RB Olsen; *Sorlandet Hospital, Kristiansand* M Haugli; *Stavanger University Hospital* B Berg; *Telemark Hospital* HK Skudal; *Trondheim University Hospital* R Hannula; *University Hospital of North Norway* AB Kildal; *Vesfold Hospital* A Johannessen; *Vestre Viken Hospital Trust, Bærum* AA Tveita; *Vestre Viken Hospital Trust, Drammen* L Heggelund; *Vestre Viken Hospital Trust, Kongsberg* G Ernst; *Vestre Viken Hospital Trust, Ringerike* L Thoresen.

**Pakistan:** *Agha Khan University Hospital, Karachi* D Begum, F Mahmood, N Nasir; *Pakistan Institute of Medical Sciences, Islamabad* N Akhtar, U Walayat; *Shaukat Khanum Memorial Cancer Hospital and Research Centre* A Raza; *The Indus Hospital, Karachi* S Bhatti, F Herekar, M Hussain, S Mustafa, A Rahim, A Rehman, S Sarfaraz, Q Shaikh.

**Peru:** *Centro Médico Naval* KH Bernal-Málaga, KCM Del-Aguila-Torres, DY Gastiaburú-Rodriguez, AA Gomero-Lopez, M Laca-Barrera, CX Peña-Mayorga, J Pro, JM Samanez-Pérez, GM Sotomayor-Woolcott; *Clínica Ricardo Palma* GE Gianella-Malca, OJ Ponce, KM Rojas-Murrugarra, RKA Tapia-Orihuela; *Clínica San Pablo* CV Luna-Wilson, FJ Ortega-Monasterios, A Peña-Villalobos; *Hospital Cayetano Heredia* CR Cornejo-Valdivia, G Málaga, F Mejía-Cordero; *Hospital de la Amistad Perú Corea Santa Rosa II* JA Juárez-Eyzaguirre, F León-Jiménez; *Hospital III Daniel Alcides Carrión-ESSALUD* LG Barreto-Rocchetti, N Flores-Valdez, MA Hueda-Zavaleta, MA Inquilla-Castillo, JA Mendoza-Laredo, JP Otazú-Ybáñez, KE Ponte-Fernandez, OJ Vargas-Anahua; *Hospital María Auxiliadora* AM Alva-Correa, B Ángeles-Padilla, RA Franco-Vásquez, RC Gallegos-López, M Olivera-Chaupis, MA Paredes-Moreno, W Torres-Ninapayta, RD Vásquez-Becerra; *Hospital Nacional Alberto Sabogal Sologuren* EC Agurto-Lescano, LE Hercilla-Vásquez, CA Iberico-Barrera, CS Terrazas-Obregón; *Hospital Nacional Daniel Alcides Carrión* J Castillo-Espinoza, JN Chacaltana-Huarcaya, E Díaz-Chipana, CM Quispe-Nolazco, ME Ramos-Samanez, JG Vásquez-Cerro, RM Yauri-Lazo; *Hospital Nacional Dos de Mayo* HC Arbañil-Huamán, CV Ibarcena-Llerena, GF Miranda-Manrique, G Santos-Revilla, VF Terrones-Levano, CE Ticona-Huaroto, DY Ugarte-Mercado; *Hospital Nacional Hipólito Unanue* A Soto; *Hospital Nacional Hipólito Unanue* AM Alcantara-Díaz, JA Azañero-Haro, RJ Carazas-Chavarry, A Cruz-Chereque, RM Sánchez-Sevillano; *Hospital Nacional Sergio E. Bernales* IC Casimiro-Porras, ODC Peña-Vásquez, E Sánchez-Garavito, H Sandoval-Manrique, JA Silva-Ramos, OM Torres-Ruiz; *Hospital Regional Lambayeque* ED Meregildo-Rodríguez; *Hospital Regional Lambayeque* JG Alvarado-Moreno, PC Ávila-Reyes, JMA Benitez-Peche, LN Cabrera-Portillo, HC Sánchez-Carrillo, MA Solano-Ico, M Villegas-Chiroque; *Universidad Peruana Cayetano Heredia* PM Cárcamo, AL Williams).

**Philippines:** *Asian Hospital and Medical Center* L Fernandez, M Kwek; *Baguio General Hospital* TPT Cajulao; *Batangas Medical Center* RJ Javier; *Cardinal Santos Medical Center* MSA Ramos, LEG Santos; *Cebu Doctors' University Hospital* MM Chua, G Garcia; *Chinese General Hospital* KL Li; *Diliman Doctors Hospital* GM Europa, D Tagarda; *Fe Del Mundo Medical Center* KL Ngo-Sanchez; *Lung Center of the Philippines* V De los Reyes, MC Orden; *Makati Medical Center* J Caoili, MT Gler; *Manila Doctors Hospital* SMA Andales-Bacolcol, MJ Nepomuceno, D Teo; *ManilaMed, Medical Center Manila* EA Roxas, BM Te; *Perpetual Succor Hospital, Cebu* P Blanco, MB Chua, MC Mujeres; *Research Institute for Tropical Medicine* JU Garcia, ADE Roman; *San Juan de Dios Educational Foundation Hospital* RD Paez, C Ramos; *San Lazaro Hospital* JT Arches, AG Awing, R Solante, DR Ymbong; *Southern Philippines Medical Center* I Chin, A Lee, K Roa; *St. Luke's Medical Center Global* M Panaligan; *St. Lukes Medical Center Quezon City* RM Llorin, JMA Quinivista-Yoon, JJ Suaco, CJ Tibayan; *St. Luke's Medical Center Quezon City* GMA Zabat; *The Medical City* CLR Abad, EA Aventura, J Bello, J Francisco, MA Lansang; *University of the East Ramon Magsaysay Memorial Medical Center* J Cabrera, V Catambing, MC Rosario; *University of the Philippines, Philippine General Hospital* MS Arcegono, SV Buno, A David-Wang, AF Malundo, RE Villalobos; *Vicente Sotto Memorial Medical Center* MV Bala, OK Macadato; *World Citi Medical Center* SM Reyes, IR Tang.

**Saudi Arabia**: *Al Noor Specialist Hospital Mekkah* M Al Gethamy, A Naji; *Dammam Central Hospital* MS AL-Mulaify; *King Faisal Specialist Hospital and Research Centre, Riyadh* A Alrajhi, R Al Maghraby; *King Khaled University Hospital, Riyadh* N Alotaibi, F AlShaharani, A Al Sharidi, M Barry, L Ghonem; *Ohud Hospital Al Madinah* A Khalel AM Kharaba; *Prince Mohammed Bin Abdulaziz Hospital, Riyadh* L Alabdan, MS AlAbdullah; *Qatif Central Hospital* A Al Shabib.

**South Africa***:* *Chris Hani Baragwanath Academic Hospital* C Menezes, SA van Blydenstein, M Venter; *Groote Schuur Hospital* M Mendelson, B Sossen; *Sefako Makgatho Health Sciences University* VL Maluleke, AN Mdladla, M Nchabeleng; *Wits Health Consortium University of the Witwatersrand* J Bennet, N Mbhele, N Mwelase, V Parker, M Rassool; *Wits Reproductive Health and HIV Institute* T Palanee-Phillips.

**Spain:** *Complejo Asistencial de Segovia* EM Ferreira Pasos; *Complejo Hospitalario de Toledo* J González Moraleja, MP Toledano; *Hospital Clínic-IDIBAPS, University of Barcelona, Barcelona* A Carrillo, M Chumbita, L De la Mora, F Etcheverry, F Garcia, M Hernández, A Inciarte, L Leal, O Miró, A Moreno, P Puerta, M Solà, A Soriano, A Tomé; *Hospital Clinico San Carlos, UCM, IdISSC, Madrid* A Ascaso, I Burruezo, N Cabello-Clotet, V Estrada, A Leone, D Lozano-Martin, FJ Martín, MJ Nuñez Orantos, AB Rivas Paterna, I Sagastagoitia, R Sandoval, E Vargas; *Hospital Clínico Universitario Lozano Blesa IIS Aragón, Zaragoza* MJ Esquillor-Rodrigo, J Guzmán, JR Paño-Pardo, C Toyas-Miazza; *Hospital Comarcal de Blanes* E Torres García; *Hospital Comarcal Sant Jaume de Calella* J Algarra Vento, O Del Rio Pérez, A Macias Paredes, D Pelleja Munné, S Valero Rovira; *Hospital Consorcio General Universitario Valencia* M García Deltoro, P Ortega, F Puchades, F Sanz, J Tamarit; *Hospital de Manises* K Jerusalem; *Hospital de Mérida* AM Pérez Fernández; *Hospital General de Tomelloso* MI Elices-Calzón, J González-Cervera, G López-Larramona, AJ Lucendo, MM Maestre-Muñiz, M Martín-Toledano, S Masegosa-Casanova, AM Ruiz-Chicote; *Hospital General Universitario de Alicante* I Agea, V Boix, R García, J Gil, P Llorens, E Merino, S Reus, R Sánchez, D Torrús-Tendero; *Hospital General Universitario de Elche, Alicante* F Gutierrez, M Masiá , S Padilla; *Hospital General Universitario Gregorio Marañón* J Berenguer, P Diez, C Diez, C Fanciulli, I Gutiérrez, I Miguens, L Pérez-Latorre, M Ramirez; *Hospital La Paz.IdIPAZ* JR Arribas, F de la Calle, B Díaz Pollán, MR Torres; *Hospital Puerta de Hierro* GAC Adolfo Centeno, AC Caballero, ADS Diaz De Santiago, AFC Fernandez Cruz, EMR Muñez Rubio, IPP Pintos Pascual, ARM Ramos Martinez; *Hospital Quirón Salud Córdoba* A Juan Arribas; *Hospital Regional Universitario de Malaga* R Gomez-Huelgas, MD Lopez-Carmona, I Perez-Camacho; *Hospital*

*Universitari Sagrat Cor* R Salas; *Hospital Universitario Araba* JC Gainzarain, MA Moran, Z Ortiz De Zarate, J Portu, E Saez De Adana; *Hospital Universitario Basurto* JM Baraiaetxaburu Artetxe, M De La Peña Trigueros, J De Miguel Landiribar, OL Ferrero Beneitez, S Ibarra Ugarte, M Intxausti Urrutibeaskoa, I Lombide Aguirre, I Lopez Azkarreta, M López Martínez, P Muñoz Sanchez, V Polo San Ricardo, A Sagarna Aguirrezabala, MZ Zubero Sulibarria; *Hospital Universitario de Badajoz* FF Rodríguez Vidigal; *Hospital Universitario de Ceuta* D García Muñiz, E Laza Laza, M Sangüesa Jareño; *Hospital Universitario de Cruces* A Basterretxea Ozamiz, MJ Blanco Vidal, M Del Alamo Martinez, A García de Vicuña Melendez, AJ Goikoetxea Agirre, M Ibarrola Hierro, I Isasi Otaolea, J Nieto Arana; *Hospital Universitario de Getafe* DAP Abad Pérez, EAR Aranda Rife, MBR Balado Rico, ECS Conde Senovilla, MCS Del Cerro Saélices, LFO Fernández de Orueta, AHR Herrera Rodríguez, NLP López Muñoz, MLL Luengo López, EM Manzone, BMC Martínez Cifre, MMF Muñoz Flores, SOS Odeh Santana, GPC Pérez Caballero; *Hospital Universitario de Jaén* C Alarcón-Payer, MJ Barbero Hernández, C Herrero Rodríguez, F Horno Ureña, FJ La Rosa Salas, G Pérez Chica; *Hospital Universitario de Salamanca* JA Martín Oterino; *Hospital Universitario Donostia* E Agirre, I Alvarez, A Berroeta, MJ Bustinduy, X Camino, A Couto, A Fuertes, MA Goenaga, M Ibarguren, JA Iribarren, X Kortajarena, MA Von Wichmann, JJ Zubeldia, B Zubeltzu, A Zufiaurre; *Hospital Universitario Fundación Alcorcón* MA Abreu-Galan, O Devora-Ruano, M Galan de Juana, C Guijarro, J Hernandez-Nuñez, JJ Martínez-Simón, O Martin-Segarra, A Pablo-Esteban, JM Parra-Ramirez, JT Pérez-Hopkins, MP Pozo-Peña, G Sierra-Torres, A Vegas-Serrano, M Velasco; *Hospital Universitario Infanta Leonor* EA Alvaro-Alonso, P Ryan, J Valencia; *Hospital Universitario Infanta Sofía* P Ruiz-Seco; *Hospital Universitario Río Hortega de Valladolid* J Gómez Barquero; *Hospital Universitario San Pedro de Alcántara* JFMJ Juan F. Masa; *Hospital Universitario Son Espases* J Asensio, F Fanjul, A Ferre, M I Fullana, M Peñaranda, L Ramon; *Hospital Universitario Virgen de la Victoria, Málaga* R Jiménez-López, E Nuño, C Pérez-López, J Sánchez-Lora, E Sánchez-Yáñez; *Hospital Universitario Virgen Macarena* MD Del Toro, M Gutiérrez-Moreno, I Jiménez-Varo, Z Palacios-Baena, N Palazón-Carrión, P Retamar, J Rodríguez-Baño, E Salamanca-Rivera, M Sevillano, A Valiente-Méndez, D Vicente-Baz; *Hospital Universitario y Politécnico La Fe, Valencia* PBG Pablo Berrocal Gil, M Salavert Lletí; *Hospital Universitario12 de Octubre, Madrid* A Lalueza IIS Aragón M de la Rica, L Diez-Galán; *Ramón y Cajal Hospital, Madrid* Y Aranda García , P Borque, S Chamorro Tojeiro , B Comeche, N Diaz Garcia, R Escudero-Sanchez, F Gioia, B Monge-Maillo, S Moreno Guillen, R Ron Gonzalez, P Vizcarra.

**Switzerland:** *Campus SLB, Lindenhofgruppe Bern* A Bosshard, J Wiegand; *Clinic of Infectiology and Infection Control, Kantonsspital Baden* M Greiner; *Department of Internal Medicine, Kantonsspital Frauenfeld* S Gastberger; *Hôpital du Valais Sion* N Desbaillets, S Emonet, PA Petignat, E Stavropoulou; *Hôpital fribourgeois Fribourg* V Erard; *Hôpital Riviera-Chablais Rennaz* F Duss, N Garin; *Hôpitaux universitaires de Genève* A Calmy, Y Flammer, A Marinosci, V Prendki; *Kantonsspital Aarau* A Conen, S Haubitz, B Jakopp, E West; *Kantonsspital Baden* B Wiggli; *Lausanne University Hospital* F Desgranges, D Haefliger, V Suttels, L van den Bogaart; *Réseau hospitalier neuchâtelois Neuchâtel* O Clerc; *Spital Thurgau AG, Kantonsspital Münsterlingen* R Fulchini, *Universitätsspital Basel* M Stoeckle.

#### Data management and monitoring

**Castor EDC, The Netherlands:** D Art, E Kochova.

**University of Bern, Switzerland:** S Trelle, S McGinty, M Branca, S Appadoo

**University of Bristol, United Kingdom:** JAC Sterne, CA Rogers, HBC Cappel-Porter, D Hutton, S Bellani, E Allum, J Kirwan.

**University of Oxford, United Kingdom:** HC Pan.

**WHO HQ, Switzerland:** P Boucher, G Vasilache

#### Statistical analysis

**University of Oxford, United Kingdom:** HC Pan, R Peto.

#### WHO trial coordination team

AM Henao-Restrepo, P Lydon, MC Miranda-Montoya, M-P Preziosi, K Salami K, V Sathiyamoorthy, S Swaminathan.

#### Other collaborators in participating countries

**Special** recognition to all the research staff and medical teams in each of the participating hospitals, in **Argentina**: Hospital Ramos Mejía Buenos Aires (SA Arrigorriaga, RF Fernandez Deu, AG Guida), and Hospital Rawson Córdoba (LK Lassen, SF Silva, CT Toledo, AZ Zamora, LZ Zappia); **Austria**: AGMT Arbeitsgemeinschaft Medikamentöse Tumortherapie, Salzsburg (S Esmaeilzadeh-leithner, B Lamprecht, D Wolkersdorfer); **Belgium**: Cliniques universitaires de Bruxelles (Z Khalil) **Brazil:** Ministry of Health (CG Sachetti, FF Soares), Hospital Universitário Clementino Fraga Filho (RA Medronho), Hospital Estadual de Sumaré (MJ Moraes), Oswaldo Cruz Foundation (BGJ Grinsztejn, MA Krieger), Hospital Hospital Federal do Rio de Janeiro (ALM Oliveira), Hospital Regional de São José (M Vieira), and Hospital São José de Doenças Infecciosas (CFV Takeda); **Canada**: Hôpital Charles-Le Moyne (G Poirier), Dr Evert Chalmers Hospital (Z Aslam), Montfort Hospital (N Chagnon), Centre Hospitalier de l'Université de Montréal (M Durand, S Matte), Hôpital du Sacré-Cœur de Montréal (YA Cavayas), Saint Paul’s Hospital Vancouver (W Connora, N Press), Sunnybrook Hospital (P KIIza, E Shadowitz), University of Alberta (A Singh), University of British Columbia (V Chaubey, J Grant, A Mah), University Health Network Toronto (B Coburn, SM Poutanen), University of Manitoba (A Heendeniya, LE Kelly), McMaster University (J Tsang), and Unity Health Toronto (KL Schwartz); **Colombia**: Clínica Reina Sofia, (M Choconta, L Martínez), Clínica Universitaria Colombia (Y Gil, M Jiménez, A Montañez, O Córdoba), Clínica Santa María del Lago (O Agudelo, J de La Hoz, M Salazar, A Valencia), Clínica Sebastián de Belalcázar (A Muriel, J Villabon), Clínica iberoamerica (C Arévalo, C Rebolledo), Fundación Cardio Infantil-Instituto del Corazón (L Sáenz, J Villar), Fundación Santa fe de Bogotá (S Bello), Fundación Valle del Lili (K Gómez, A Martínez, A Sotomayor, J Yara), Fundación Universitaria Sanitas (C Aristizábal, D Castro, M Isaza, P Marín, C Orjuela), Hospital Universitario San Ignacio (V Méndez, C Gómez), Hospital Universidad del Norte (S Aguilera, F Torres), Ministry of Health (A Moscoso, F Ruiz), PAHO (L. Ramírez, G Tambini), INVIMA (J Aldana, P Pulgarín); National University of Colombia (M Jimenez, O Cordoba, A Montañez); **Finland**: Helsinki University Hospital (P Järvinen, I Kalliala, TP Kilpeläinen), Occupational Health Helsinki (JMJ Mustonen), and Tampere University Hospital (RH Hankkio, GM Määttä, VK Virtanen); **France:** Hôpital Avicenne Paris (O Bouchad),

Hôpital d'Instruction des Armées Bégin (C Ficko), Hôpital Bichat Paris (B Basli, A Chair, J Level, M Schneider, J Guedj, C Laouenan, S Tubiana, V Godard), Hôpital de Bicêtre (X Monnet), Hospices Civils de Lyon (B Leveau), Centre hospitalier universitaire de Martinique (A Cabie), Pontchaillou University Hospital (M Revest), Reims University Hospital (F Bani-Sadr, Rennes University Hospital (A Caro, C Cameli, MJ Ngo Um Tegue); Nouvel Hôpital Civil Strasbourg (JL Diehl), Tours University Hospital (L Bernard), ANRS (V Petrov-Sanchez, S Le Mestre, C Cagnot, D Lebrasseur, C Birkle, C Moins, S Gibowski, C Paul, E Landry, E Balssa, L Wadouachi, A le Goff, L Moachon), ANSES (C Semaille), Imagine Institute Paris (L Abel), Inserm (H Esperou, S Couffin-Cadiergues, E d'Ortenzio, B Hamze, O Puechal), Inserm ANRS Villejuif (Y Riault, E Netzer), Infective Agents Institute Lyon (M Bouscambert-Duchamp, V Icard, B Lina, F Morfin-Sherpa, A Gaymard), Université Bordeaux Inserm (L Wittkop, L Moinot, A Gelley), Université Paris Saclay Inserm (A Essat, M Ghislain, M Brossard), and Université Sorbonne Inserm (L Beniguel, M Genin); **Honduras**: Hospital Atlantida la Ceiba (M Juarez); Instituto Cardiopulmonar Tegucigalpa (N Maradiaga), Hospital Leonardo Martinez San Pedro Sula (J Samara), Hospital Militar Tegucigalpa (JS Jerez), Hospital San Felipe Tegucigalpa (E Cruz, H Rodriguez), Agencia de Regulación Sanitaria (F Contreras), National Autonomous University of Honduras (F Herrera, S Moncada, W Murillo), Secretaria de Salud de Honduras (A Flores, R Aplicano), and PAHO (P Huerta); **India:** SVP Institute of Medical Sciences and Research Ahmedabad (N Suthar, S Shah, P Palat, A Chandwani, A Pandya,V Buch, S Talati, D Patel), AIIMS Bhopal (V Ingle, A Singhai, N Shrivastava), Apollo Hospitals Greams Lane Chennai (S Pavithra, E Elvira, G Parthasarathy, Y Arun Chander, A Afsal, NK Hilda), Apollo Speciality Hospitals Vanagaram Chennai (J Krishnan, S Hilda, K Kirubanandam, C Poongavanam, J Swaminathan), Madras Medical College Chennai (G Arathi, G Jayashree, T Meenakshi, S Gomathi), Omandurar Medical College Chennai (C R Anuradha, R Pravin Kumar, S Sai Vishal, C Praveen Kumar), VHS Infectious Diseases Medical Centre Chennai (F Beulah, S Ramu, G Narayanan), Gandhi Hospital Hyderabad (B Sheshadri, MD Iqbal Ahmed), SMS Medical College & Hospital Jaipur (B Goyal), BYL Nair Hospital Mumbai (R Singh, A Bhamare), PD Hinduja National Hospital and Medical Research Centre Mumbai (A Sunavala, S Mehendale, RS Raju), Government Medical College Nagpur (M Faisal, P Gomase, P Gosavi, S Bhelekar, P Agrawal, N Agrawal, R Sabu), AIIMS New Delhi (R Subramaniam, A Anant, S Bhatnagar, L Dar, S Bhoi, P Mathur, A Kumar, M Ved Prakash, P Tiwari), Indian Council of Medical Research New Delhi (M Murhekar, S Agrawal, B John), the Army Institute of Cardio Thoracic Sciences Pune (V Mangal), Bharati hospital Pune (S Palkar), ICMR- National AIDS Research Institute Pune (S Krishnan, R Bangar, P Kerkar, K Chaudhari, P Kokate, A Kashikar), AIIMS Rishikesh (M Singh, A Chauhan), Government Medical College Surat (A Patel, K Chauhan), and Christian Medical College Vellore (T George, L Audrin, G Karthik, GM Varghese, P Rupali, T Balamugesh, V Surekha, B Chacko, M Moorthy, K P P Abhilash, SC Nair, S Chandy, R Charles, A Jacob, D Mathew, E Inbarani, R Moses, N Stanely); **Lebanon**: Ministry of Public Health (R Hamra); **Lithuania**: Hospital Santaros klinikos, Vilnius, (M Paulauskas, U Sakalauskiene) **Luxembourg**: Clinical and Epidemiological Investigation Center, Strassen (M Alexandre), Hôpitaux Robert Schuman, Luxembourg (M Berna) **Pakistan**: Hayatabad Medical Complex Peshawar (S Jamal), Shaukat Khanum Memorial Cancer Hospital (S Hassan, S Abbas, S Khan, MR Khan), and Shifa International Hospital (E Khan, S Azam); **Philippines**: Asian Hospital and Medical Center (MIL Fernandez), Baguio General Hospital (MLF de Leon, RG Dagwasi), Batangas Medical Center (ML Almero, M Mercado), Cardinal Santos Medical Center (A Vergara), Cebu Doctors' University Hospital (F Repunte), Chinese General Hospital (SO Tan, MK Ong-Tantuco, RC Reyes, PLG Co, ALG Gabriel-Chan, AO Reyes-Addatu, JT Li-Yu, SA Ang), Diliman Doctors Hospital (KI Del Ayre), Fe Del Mundo Medical Center (SMA Santos, A Torrico), Lung Center of the Philippines (H Basobas, Z Del rosario), Makati Medical Center (KM Taladua, HF Ricaforte-Docuyanan), Manila Doctors Hospital (A Ramos-Precilla, SA Limson), Medical Center Manila (TAE Nunez), The Medical City (A Santiago), Perpetual Succor Hospital, Cebu (TR Cuevas), Philippine Council for Health Research and Development (JC

Montoya ), Philippine General Hospital (JP Benedicto, M Llanes, G Astudillo, P Nala, R Abaya), Research Institute for Tropical Medicine (A Yabut), San Juan de Dios Educational Foundation Hospital (JD Cruz), San Lazaro Hospital (SM Ligutan), St. Lukes Medical Center Quezon City (AR Cumpas), Vicente Sotto Memorial Medical Center (M Bagano, MP Pablo-Villamor, GM Aquino Jr, JD Bancat), Southern Philippines Medical Center (MYC Barez, LL Torno, P Ferrer, EA Sibal),St. Lukes Medical Center Global (G Dy-Arga, R Enecilla, CE Villavicencio, DD Ona, S Unson, JD Gargar, BM Samonte), University of the East Ramon Magsaysay Memorial Medical Center (MTF Sumagaysay), University of the Philippines (FM Climacosa, ME Mercado, CDA Rozul, P Tagle, M Recana), World Citi Medical Center (DJD Reotita), Philippine Clinical Research Professionals (G Mendoza, J Arellano, AR Baniqued), Department of Health Philippines (FT Duque III, MR Vergeire), Food and Drug Administration Philippines (RE Domingo), and WHO WPRO (JP Tonolete); **Saudi Arabia:** King Faisal Specialist Hospital and Research Centre, Riyadh (N Alorayyidh, R Moslmani), King Khaled University Hospital, Riyadh (A Abdurrahman, D Bintaleb); **Spain:** Hospital Clinico San Carlos, UCM, IdISSC, Madrid (V Alvarez, C Perez-Ingidua, N Pérez Macias, O Bueno, MJ Tellez), and Hospital La Paz Madrid (A Borobia); **South Africa**: Chris Hani Baragwanath Academic Hospital (D Kalambay, WBT Lechuti), Groote Schuur Hospital (S Koekemoer, S Moosa, T Morar), Sefako Makgatho Health Sciences University (SC Shaku, VM Ramothwala), Wits Health Consortium (C Barker, J Chelliah, J Ferreira, M Knight, LI Koeberg, Y Kilian, A Rama, D Strydom, S Naidoo), and Wits Reproductive Health and HIV Institute (F Docrat, A Jacques, K Moodley, T Msomi, R Boikanya, S Cornell); **Switzerland**: Kantonsspital Baden (A Friedl, J Rutishauser, F Rutz), Campus SLB Lindenhofgruppe Bern (C Groen, J Evison), Kantonsspital Frauenfeld (P Rochat, P Hackman, P Wiesli, A Kistler, R Ursprung, S Danioth, R Werner, S Dias, M Schuster), Hôpitaux universitaires de Genève (P Vazquez, Y Gosmain), Lausanne University Hospital (M Cavassini, A Fayet-Mello, L Vallotton, L Warpelin-Decrausaz, V Sormani, D Niksch), and Hôpital du Valais Sion (M Eyer, E Schaefer, S Schwery, MSavet, A Luyet); and **WHO teams**: AFRO (C Garapo, JP Okeibunor), EMRO (A Hashish, C Kodama, A Mandil), EURO (C Butu, M Dara, A Kuli, A Mesi, N Mamulashvili, I Zurlyte), PAHO/AMRO (L Reveiz), SEARO (T Azim, M Gupta, R Takahashi), WPRO (A Cawthorne, YR Lo, JP Tonolette) and Headquarters (V Benassi, S Benitez, A Borges, T Bouquet, S Chuffart, E Egorova, R Embaye, S Kone, C Merle, P Molinaro, R La Rotta, MJ Ryan, N Mafunga, A Mazur, G Queyras).

#### Acknowledgments

The chief acknowledgement is to the thousands of patients and their families who participated in this trial and made it possible, and the hundreds of medical staff who randomized and cared for patients.

The Ministries of Health of the participating Member States and their institutions provided critical support in the implementation of the trial.

Castor EDC donated and managed their cloud-based clinical data capture and management system. Anonymized data handling and analysis was at the Universities of Berne, Bristol and Oxford.

Remdesivir was donated by Gilead Sciences, Hydroxychloroquine by Mylan, Lopinavir-Ritonavir by Abbvie, Cipla and Mylan and Interferon β1a by Merck KGaA (subcutaneous) and Faron (intravenous).

Add-on studies were conducted in Canada, France, India and Norway.

### Table S1. Treatment allocation vs initiation of ventilation in those not already being ventilated at the time of randomization

Ventilation includes invasive or non-invasive mechanical ventilation or extra-corporeal membrane oxygenation.

| | Remdesivir vs its control | | Hydroxychloroquine vs its control | | Lopinavir vs its control | | Interferon vs its control* | |
|---|---|---|---|---|---|---|---|---|
| | Active | Control | Active | Control | Active | Control | Active | Control |
| **Not ventilated at entry** | **2489** | **2475** | **862** | **824** | **1287** | **1258** | **1911** | **1920** |
| Ventilated later; died | 117 | 108 | 29 | 19 | 50 | 44 | 108 | 91 |
| Ventilated later; discharged | 139 | 146 | 42 | 44 | 67 | 68 | 81 | 98 |
| Ventilated later; pending* | 39 | 30 | 4 | 3 | 7 | 7 | 20 | 21 |
| **Total ventilated (number, and %)†** | **295**<br>**11.9** | **284**<br>**11.5** | **75**<br>**8.7** | **66**<br>**8.0** | **124**<br>**9.6** | **119**<br>**9.5** | **209**<br>**10.9** | **210**<br>**10.9** |

* Ventilation can be reported in patients who have not yet died or been discharged.

† More complete follow-up will increase the numbers known to have been ventilated or died.

### Table S2. Use of corticosteroids and other non-study drugs

Numbers and percentages are tabulated

.

| | Remdesivir vs its control | | Hydroxychloroquine vs its control | | Lopinavir vs its control | | Interferon vs its control* | |
|---|---|---|---|---|---|---|---|---|
| Corticosteroids<br>Number & percentage | 1310<br>47.8 | 1288<br>47.6 | 140<br>14.8 | 140<br>15.5 | 316<br>22.6 | 328<br>23.9 | 981<br>47.9 | 1053<br>51.4 |
| Convalescent plasma | 52<br>1.9 | 58<br>2.1 | 7<br>0.7 | 3<br>0.3 | 24<br>1.7 | 15<br>1.1 | 43<br>2.1 | 33<br>1.6 |
| Anti-IL-6 drug | 133<br>4.9 | 143<br>5.3 | 21<br>2.2 | 18<br>2.0 | 42<br>3.0 | 42<br>3.1 | 52<br>2.5 | 68<br>3.3 |
| Non-trial interferon | 3<br>0.1 | 25<br>0.9 | 2<br>0.2 | 1<br>0.1 | 4<br>0.3 | 0<br>0.0 | 1<br>0.1 | 26<br>1.3 |
| Non-trial antiviral | 65<br>2.4 | 152<br>5.6 | 62<br>6.6 | 54<br>6.0 | 86<br>6.2 | 90<br>6.6 | 102<br>5.0 | 144<br>7.0 |
| Number entered | 2743<br>100.0 | 2708<br>100.0 | 947<br>100.0 | 906<br>100.0 | 1399<br>100.0 | 1372<br>100.0 | 2050<br>100.0 | 2050<br>100.0 |

#### Table S3. Multivariate analysis simultaneously estimating all 4 effects

The pre-planned primary analyses in the main text involved 4 pairwise comparisons, one between each treatment group and its controls, as indicated in the flowchart (Figure 1). These 4 primary analyses were stratified by age and by whether the patient was already ventilated at the time of randomization, and found no definitely favourable or definitely unfavourable effect of any of the 4 study drugs on all-cause in-hospital mortality (Figure 3). The RRs in these 4 pre-planned pairwise comparisons were:

Remdesivir vs its control (pre-planned analysis) RR=0.95 (95% CI 0.81-1.11; P=0.50),

Hydroxychloroquine vs its control (pre-planned analysis) RR=1.19 (0.89-1.59; P=0.23),

Lopinavir vs its control (pre-planned analysis) RR=1.00 (0.79-1.25; P=0.97), and

Interferon vs its control (pre-planned analysis) RR=1.16 (0.96-1.39; P=0.11).

As there was some overlap between the 4 control groups, an exploratory sensitivity analysis used multivariate Cox regression to fit all 4 treatment effects simultaneously, assuming the independence of any effects of Lopinavir and of Interferon. This multivariate analysis was stratified by the set of study drugs that was locally available at randomization (13 occupied strata) and was adjusted for several of the prognostic factors listed in Table 1: age (<40, 40-49, 50-59, 60-69, 70-79, 80+ years), sex, diabetes, bilateral lung lesions at entry (no, yes, not imaged at entry), and respiratory support at entry (no oxygen, oxygen but no ventilation, ventilation). This multivariate sensitivity analysis had not been pre-planned as a primary or a secondary analysis. For each of the 4 study drugs it yielded mortality rate ratios (RRs) for active treatment vs local standard of care (SoC) that were similar to those in the pre-planned primary pairwise comparisons, again finding no definitely favourable or unfavourable effect of any of the 4 study drugs:

Remdesivir vs local SoC (in multivariate analysis) RR=0.95 (95% CI 0.81-1.11; P=0.49),

Hydroxychloroquine vs local SoC (in multivariate analysis) RR=1.14 (0.89-1.46; P=0.31),

Lopinavir vs local SoC (in multivariate analysis) RR=0.94 (0.76-1.16; P=0.56), and

Interferon vs local SoC (in multivariate analysis) RR=1.14 (0.96-1.35; P=0.13).

**Figure S1. Effects on 28-day in-hospital mortality of (a) Remdesivir, (b) Hydroxychloroquine, (c) Lopinavir, (d) Interferon**

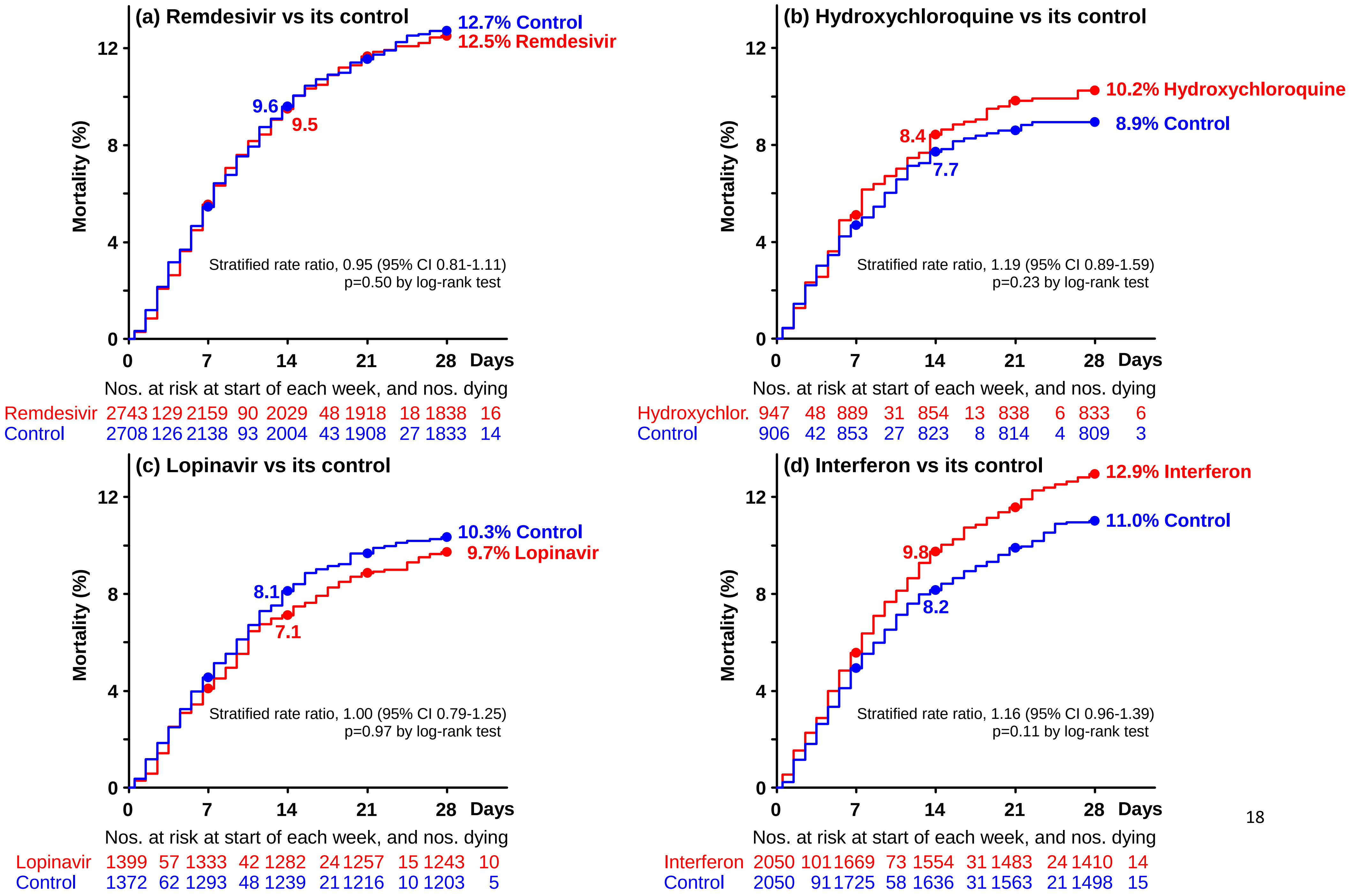

**Figure S2. Subdivision by ventilation at randomization on the apparent effects of Remdesivir on the 28-day probability of death from any cause**

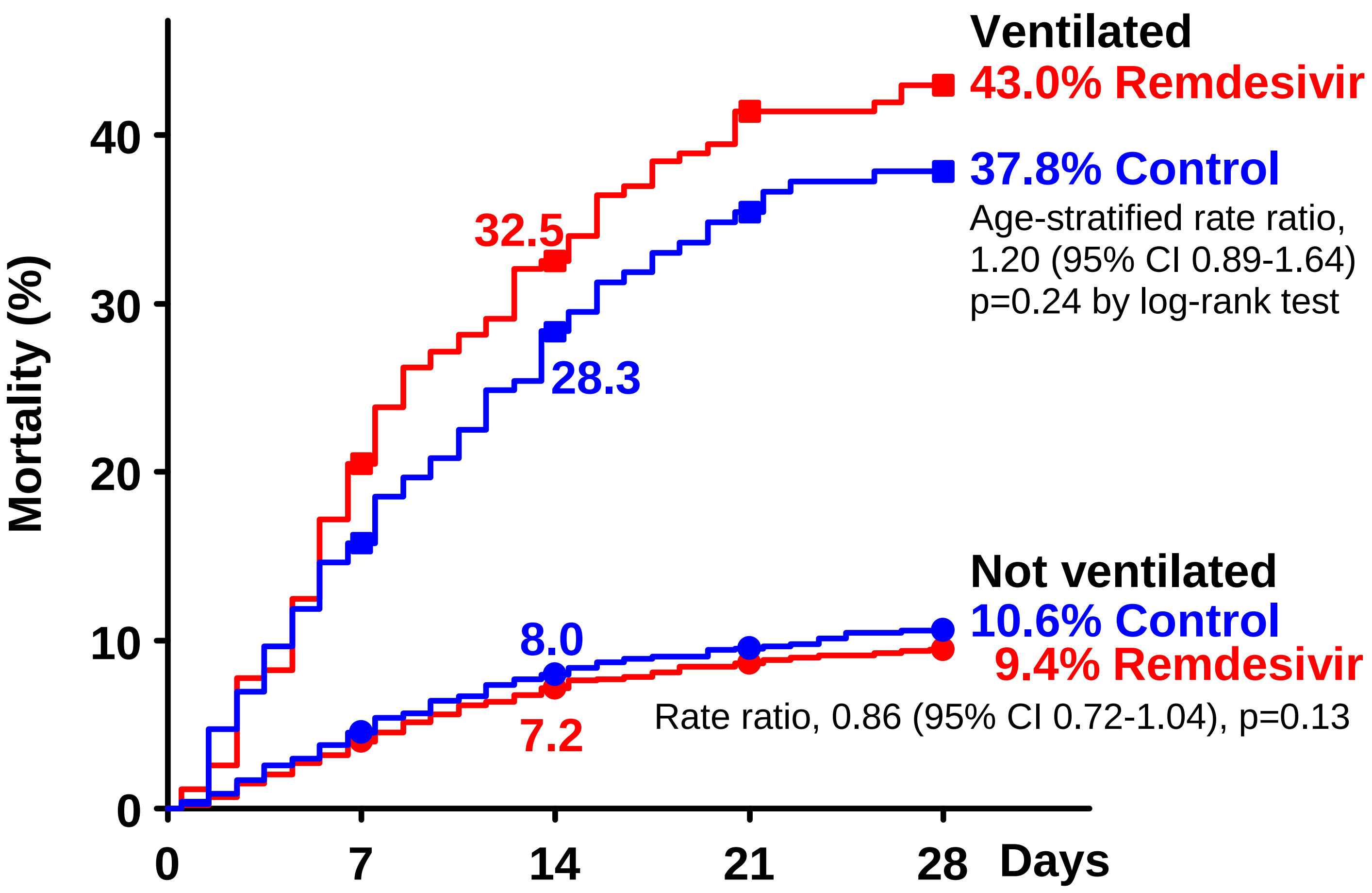


Nos. at risk at the start of each week, and nos. dying

| | | | | | | | | | | | |
|---|---|---|---|---|---|---|---|---|---|---|---|
| Remdesivir | 254 | 44 | 167 | 25 | 138 | 18 | 118 | 3 | 112 | 8 | Ventilated |
| Control | 233 | 29 | 151 | 22 | 123 | 12 | 108 | 5 | 102 | 3 | |
| Remdesivir | 2489 | 85 | 1992 | 65 | 1891 | 30 | 1800 | 15 | 1726 | 8 | Not ventilated |
| Control | 2475 | 97 | 1987 | 71 | 1881 | 31 | 1800 | 22 | 1731 | 11 | |

**Figure S3. Subdivision by ventilation at randomization on the apparent effects of Hydroxychloroquine on the 28-day probability of death from any cause**

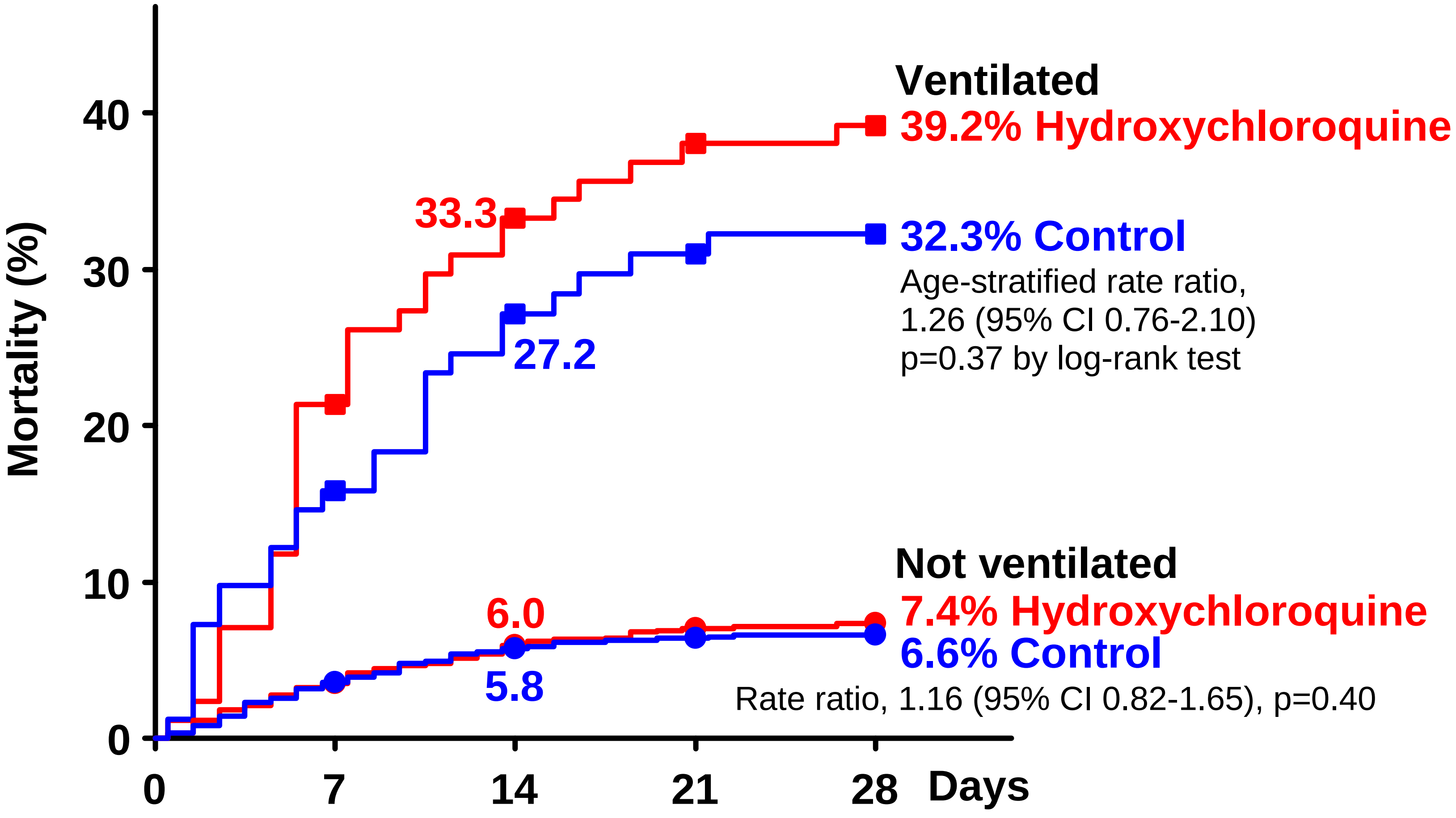


Nos. at risk at the start of each week, and nos. dying

| | | | | | | | | | | | |
|---|---|---|---|---|---|---|---|---|---|---|---|
| Hydroxyc . | 85 | 18 | 66 | 10 | 56 | 4 | 52 | 2 | 50 | 1 | Ventilated |
| Control | 82 | 13 | 69 | 9 | 57 | 3 | 54 | 2 | 53 | 0 | |
| Hydroxyc . | 862 | 30 | 823 | 21 | 798 | 9 | 786 | 4 | 783 | 5 | Not ventilated |
| Control | 824 | 29 | 784 | 18 | 766 | 5 | 760 | 2 | 756 | 3 | |

**Figure S4. Subdivision by ventilation at randomization on the apparent effects of Lopinavir on the 28-day probability of death from any cause**

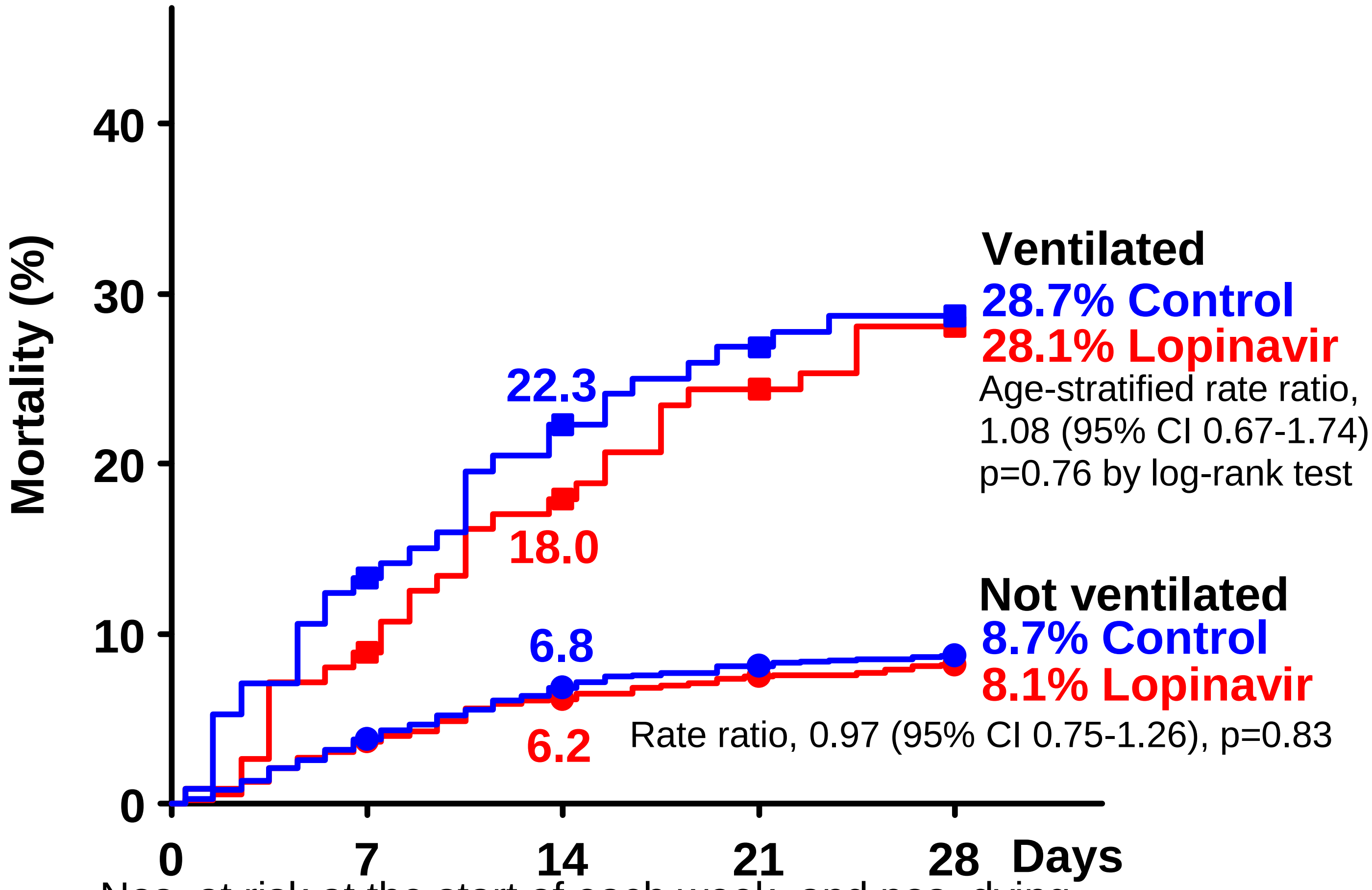


Nos. at risk at the start of each week, and nos. dying

| | | | | | | | | | | | |
|---|---|---|---|---|---|---|---|---|---|---|---|
| Lopinavir | 112 | 10 | 101 | 10 | 90 | 7 | 82 | 5 | 78 | 3 | Ventilated |
| Control | 114 | 15 | 98 | 10 | 85 | 5 | 80 | 3 | 78 | 2 | |
| Lopinavir | 1287 | 47 | 1232 | 32 | 1192 | 17 | 1175 | 10 | 1165 | 7 | Not ventilated |
| Control | 1258 | 47 | 1195 | 38 | 1154 | 16 | 1136 | 7 | 1125 | 3 | |

**Figure S5. Subdivision by ventilation at randomization on the apparent effects of Interferon on the 28-day probability of death from any cause**

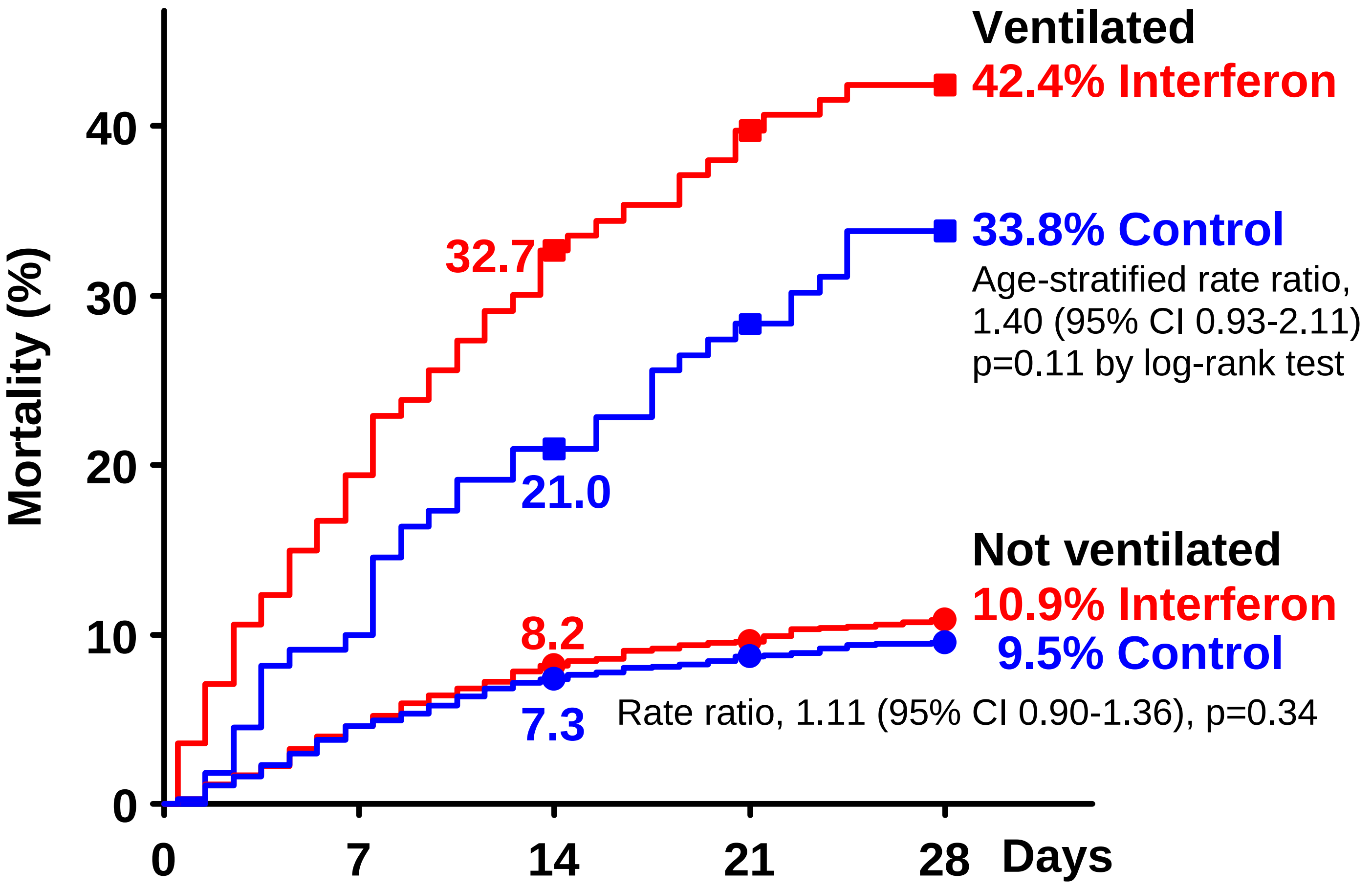


Nos. at risk at the start of each week, and nos. dying

| | | | | | | | | | | | |
|---|---|---|---|---|---|---|---|---|---|---|---|
| Interferon | 139 | 23 | 91 | 15 | 76 | 8 | 68 | 4 | 65 | 5 | Ventilated |
| Control | 130 | 11 | 99 | 12 | 86 | 8 | 78 | 6 | 72 | 3 | |
| Interferon | 1911 | 78 | 1578 | 58 | 1478 | 23 | 1415 | 20 | 1345 | 9 | Not ventilated |
| Control | 1920 | 80 | 1626 | 46 | 1550 | 23 | 1485 | 15 | 1426 | 12 | |

#### Figure S6. Rate ratios of any death, stratified by age and respiratory support at entry, Remdesivir versus Control, by entry characteristics and steroid use at any time*

| | Deaths reported / Patients randomized in ITT analyses (28-day risk, K-M%) | | Remdesivir deaths: log-rank statistics | | Ratio of death rates (RR), & 99% CI (or 95% CI, for total) |
|---|---|---|---|---|---|
| | Remdesivir | Control | O-E | Variance | Remdesivir : Control |
| **Sex** | | | | | |
| Male | 199/1706 (13.6) | 211/1725 (13.8) | -9.8 | 100.6 | 0.91 [0.70-1.17] |
| Female | 102/1037 (10.8) | 92/983 (10.8) | 2.1 | 47.9 | 1.05 [0.72-1.52] |
| **Age at entry** | | | | | |
| <50 | 61/961 (6.9) | 59/952 (6.8) | 2.3 | 29.8 | 1.08 [0.67-1.73] |
| 50-69 | 154/1282 (13.8) | 161/1287 (14.2) | -7.6 | 77.5 | 0.91 [0.68-1.21] |
| 70+ | 86/500 (20.5) | 83/469 (21.6) | -2.9 | 41.5 | 0.93 [0.63-1.39] |
| **Days from hospital admission to randomization** | | | | | |
| 0 | 79/724 (11.3) | 77/712 (11.0) | -1.3 | 38.0 | 0.97 [0.64-1.47] |
| 1 | 81/917 (10.1) | 95/938 (11.4) | -8.8 | 42.9 | 0.82 [0.55-1.21] |
| 2+ | 141/1102 (15.7) | 131/1058 (15.3) | -0.2 | 66.4 | 1.00 [0.73-1.37] |
| **Respiratory support at entry** | | | | | |
| Ventilated | 98/254 (43.0) | 71/233 (37.8) | 7.6 | 40.8 | 1.20 [0.80-1.80] |
| On oxygen | 192/1828 (12.2) | 219/1811 (13.8) | -16.9 | 101.8 | 0.85 [0.66-1.09] |
| Neither | 11/661 (2.0) | 13/664 (2.1) | -0.6 | 6.0 | 0.90 [0.31-2.58] |
| **Bilateral lung lesions** | | | | | |
| Not imaged at entry | 36/281 (15.0) | 29/296 (11.9) | 3.0 | 15.5 | 1.22 [0.63-2.34] |
| Yes | 254/2175 (13.3) | 264/2153 (13.9) | -5.4 | 127.7 | 0.96 [0.76-1.20] |
| No | 11/287 (4.5) | 10/259 (4.0) | -0.9 | 5.1 | 0.84 [0.27-2.62] |
| **Current smoking** | | | | | |
| Yes | 17/178 (10.7) | 24/161 (17.1) | -4.5 | 9.7 | 0.63 [0.28-1.43] |
| No | 284/2565 (12.6) | 279/2547 (12.4) | -3.4 | 138.4 | 0.98 [0.78-1.21] |
| **Diabetes** | | | | | |
| Yes | 98/707 (15.9) | 86/666 (14.9) | -0.6 | 44.6 | 0.99 [0.67-1.45] |
| No | 203/2036 (11.3) | 217/2042 (12.0) | -8.6 | 103.6 | 0.92 [0.71-1.19] |
| **Heart disease** | | | | | |
| Yes | 77/571 (15.3) | 75/567 (15.4) | 1.3 | 37.3 | 1.04 [0.68-1.58] |
| No | 224/2172 (11.8) | 228/2141 (12.0) | -10.0 | 110.7 | 0.91 [0.72-1.17] |
| **Chronic liver disease** | | | | | |
| Yes | 7/36 (24.4) | 6/41 (17.6) | 1.5 | 2.4 | 1.82 [0.35-9.45] |
| No | 294/2707 (12.3) | 297/2667 (12.6) | -8.9 | 145.6 | 0.94 [0.76-1.16] |
| **Chronic lung disease** | | | | | |
| Yes | 25/151 (18.2) | 19/145 (16.9) | 1.5 | 10.6 | 1.15 [0.52-2.54] |
| No | 276/2592 (12.2) | 284/2563 (12.5) | -9.7 | 138.0 | 0.93 [0.75-1.16] |
| **Asthma** | | | | | |
| Yes | 20/139 (16.2) | 15/139 (14.0) | 2.0 | 8.4 | 1.27 [0.52-3.10] |
| No | 281/2604 (12.3) | 288/2569 (12.7) | -11.0 | 140.0 | 0.92 [0.74-1.15] |
| **Corticosteroids at entry or later (ignoring any biases*)** | | | | | |
| Yes | 210/1310 (16.6) | 225/1288 (17.9) | -5.6 | 106.8 | 0.95 [0.74-1.22] |
| No | 91/1433 (8.0) | 78/1420 (6.9) | 1.2 | 41.1 | 1.03 [0.69-1.54] |
| **Geographic location** | | | | | |
| Europe or Canada | 33/715 (5.2) | 27/698 (5.1) | -1.1 | 14.5 | 0.93 [0.47-1.82] |
| Latin America | 82/470 (20.6) | 115/514 (23.8) | -4.4 | 46.2 | 0.91 [0.62-1.33] |
| Asia and Africa | 186/1558 (13.3) | 161/1496 (12.0) | 3.4 | 84.1 | 1.04 [0.79-1.38] |
| **Total** | **301/2743 (12.5)** | **303/2708 (12.7)** | **-8.3** | **148.8** | **0.95 [0.81-1.11]** **2p = 0.50** |

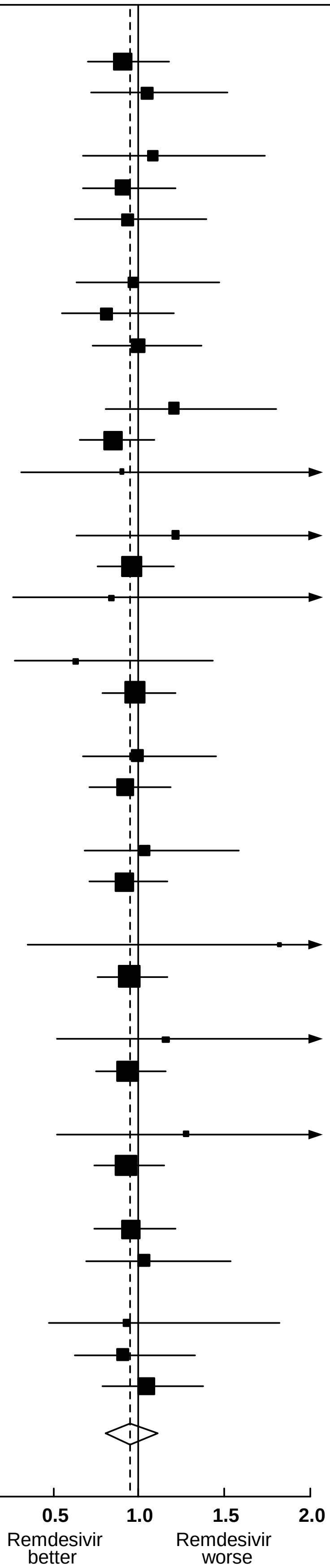


**Heterogeneity test:** $\chi^2_{18}$ **= 12.4; p = 0.82**

■ 99% or ◇ 95% confidence interval (CI), K-M Kaplan-Meier.

* RRs in steroid subgroups may be somewhat biased by later steroid use.

#### Figure S7. Rate ratios of any death, stratified by age and respiratory support at entry, Hydroxychloroquine vs. Control, by entry characteristics and steroid use at any time*

| | Deaths reported / Patients randomized in ITT analyses (28-day risk, K-M%) | | HCQ deaths: log-rank statistics | | Ratio of death rates (RR), & 99% CI (or 95% CI, for total) |
|---|---|---|---|---|---|
| | HCQ | Control | O-E | Variance | HCQ : Control |
| **Sex** | | | | | |
| Male | 80/574 (12.9) | 50/535 (9.1) | 13.4 | 31.9 | 1.52 [0.97-2.40] |
| Female | 24/373 (6.2) | 34/371 (8.7) | -5.3 | 14.2 | 0.69 [0.35-1.36] |
| **Age at entry** | | | | | |
| <50 | 19/335 (5.7) | 19/317 (5.8) | 0.9 | 9.2 | 1.10 [0.47-2.57] |
| 50-69 | 55/410 (12.1) | 31/396 (7.1) | 10.8 | 21.2 | 1.66 [0.95-2.91] |
| 70+ | 30/202 (14.0) | 34/193 (17.8) | -3.5 | 15.8 | 0.80 [0.42-1.53] |
| **Days from hospital admission to randomization** | | | | | |
| 0 | 23/296 (7.8) | 20/281 (6.8) | 0.6 | 10.6 | 1.06 [0.48-2.34] |
| 1 | 39/317 (10.8) | 29/312 (9.0) | 2.8 | 16.1 | 1.19 [0.63-2.26] |
| 2+ | 42/334 (12.0) | 35/313 (10.8) | 3.7 | 19.0 | 1.21 [0.67-2.19] |
| **Respiratory support at entry** | | | | | |
| Ventilated | 35/85 (39.2) | 27/82 (32.3) | 3.4 | 14.8 | 1.26 [0.65-2.46] |
| On oxygen | 61/517 (10.8) | 52/483 (10.5) | 2.9 | 28.1 | 1.11 [0.68-1.80] |
| Neither | 8/345 (2.3) | 5/341 (1.2) | 1.4 | 3.2 | 1.56 [0.37-6.54] |
| **Bilateral lung lesions** | | | | | |
| Not imaged at entry | 20/137 (11.9) | 18/118 (14.8) | 0.2 | 9.3 | 1.02 [0.44-2.36] |
| Yes | 78/656 (11.6) | 63/618 (9.8) | 5.9 | 34.3 | 1.19 [0.77-1.85] |
| No | 6/154 (3.3) | 3/170 (1.8) | 1.5 | 1.9 | 2.16 [0.34-13.77] |
| **Current smoking** | | | | | |
| Yes | 14/92 (14.2) | 8/82 (9.9) | 3.1 | 4.1 | 2.09 [0.59-7.41] |
| No | 90/855 (9.8) | 76/824 (8.8) | 5.3 | 40.7 | 1.14 [0.76-1.71] |
| **Diabetes** | | | | | |
| Yes | 35/199 (17.8) | 20/205 (8.3) | 8.3 | 13.2 | 1.87 [0.92-3.81] |
| No | 69/748 (8.2) | 64/701 (9.1) | 0.8 | 32.7 | 1.02 [0.65-1.61] |
| **Heart disease** | | | | | |
| Yes | 25/193 (12.0) | 20/194 (9.9) | 2.5 | 10.9 | 1.25 [0.58-2.74] |
| No | 79/754 (9.8) | 64/712 (8.7) | 5.1 | 34.9 | 1.16 [0.75-1.79] |
| **Chronic liver disease** | | | | | |
| Yes | 2/15 (6.7) | 3/14 (21.4) | -1.4 | 0.9 | 0.21 [0.01-3.14] |
| No | 102/932 (10.3) | 81/892 (8.7) | 8.9 | 45.0 | 1.22 [0.83-1.79] |
| **Chronic lung disease** | | | | | |
| Yes | 9/62 (14.7) | 7/66 (10.7) | 0.9 | 3.9 | 1.27 [0.35-4.65] |
| No | 95/885 (9.9) | 77/840 (8.8) | 6.8 | 42.2 | 1.17 [0.79-1.75] |
| **Asthma** | | | | | |
| Yes | 3/41 (7.4) | 4/46 (6.7) | 0.2 | 1.2 | 1.16 [0.11-11.85] |
| No | 101/906 (10.4) | 80/860 (9.1) | 7.8 | 44.3 | 1.19 [0.81-1.75] |
| **Corticosteroids at entry or later (ignoring any biases*)** | | | | | |
| Yes | 33/140 (23.3) | 31/140 (22.2) | 4.9 | 14.0 | 1.42 [0.71-2.83] |
| No | 71/807 (8.0) | 53/766 (6.5) | 6.5 | 30.5 | 1.24 [0.78-1.97] |
| **Geographic location** | | | | | |
| Europe or Canada | 24/286 (6.7) | 17/267 (6.1) | 3.2 | 10.1 | 1.38 [0.61-3.09] |
| Latin America | 28/97 (27.9) | 21/96 (20.8) | 4.6 | 11.4 | 1.50 [0.70-3.21] |
| Asia and Africa | 52/564 (9.0) | 46/543 (8.2) | 3.2 | 23.5 | 1.15 [0.67-1.95] |
| **Total** | **104/947 (10.2)** | **84/906 (8.9)** | **8.1** | **46.2** | **1.19 [0.89-1.59]** 2p = 0.23 |

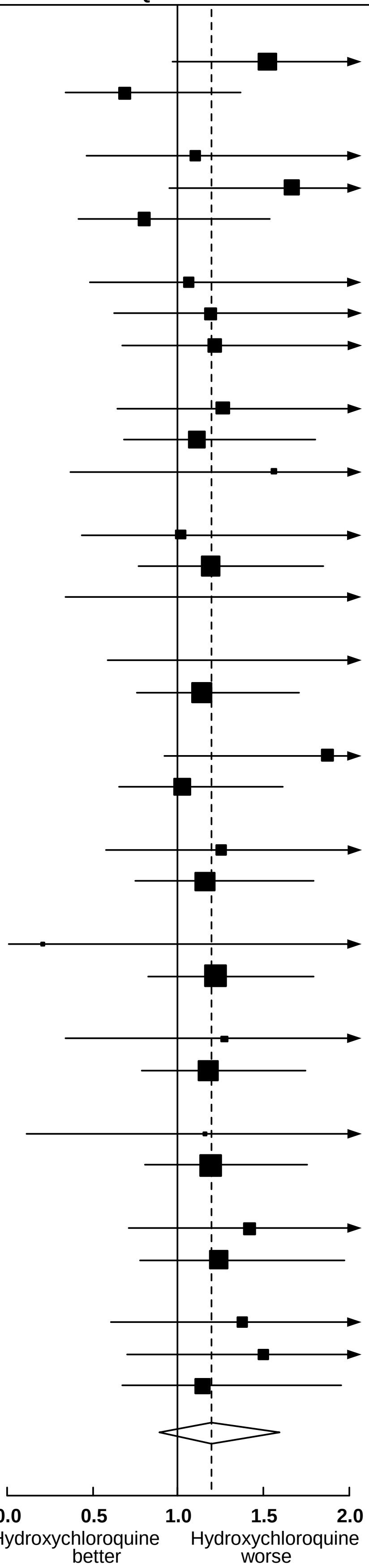


**Heterogeneity test:** $\chi^2_{18} = 21.0$; p = 0.28

■ 99% or ◇ 95% confidence interval (CI), K-M Kaplan-Meier.

* RRs in steroid subgroups may be somewhat biased by later steroid use.

#### Figure S8. Rate ratios of any death, stratified by age and respiratory support at entry, Lopinavir versus Control, by entry characteristics and steroid use at any time*

| | Deaths reported / Patients randomized in ITT analyses (28-day risk, K-M%) | | Lopinavir deaths: log-rank statistics | | Ratio of death rates (RR), & 99% CI (or 95% CI, for total) |
|---|---|---|---|---|---|
| | Lopinavir | Control | O-E | Variance | Lopinavir : Control |
| **Sex** | | | | | |
| Male | 98/851 (10.4) | 91/802 (11.0) | 1.1 | 46.3 | 1.02 [0.70-1.49] |
| Female | 50/548 (8.7) | 55/570 (9.4) | -1.8 | 25.7 | 0.93 [0.56-1.55] |
| **Age at entry** | | | | | |
| <50 | 20/511 (3.6) | 27/501 (4.9) | -3.0 | 11.7 | 0.77 [0.36-1.64] |
| 50-69 | 66/597 (9.8) | 57/596 (9.1) | 2.7 | 30.4 | 1.09 [0.68-1.74] |
| 70+ | 62/291 (20.4) | 62/275 (22.7) | 0.0 | 30.2 | 1.00 [0.63-1.60] |
| **Days from hospital admission to randomization** | | | | | |
| 0 | 34/423 (7.6) | 33/403 (7.7) | -0.8 | 16.6 | 0.95 [0.51-1.80] |
| 1 | 48/442 (9.4) | 46/445 (10.4) | 0.7 | 23.1 | 1.03 [0.60-1.76] |
| 2+ | 66/534 (11.8) | 67/524 (12.3) | 1.9 | 31.9 | 1.06 [0.67-1.67] |
| **Respiratory support at entry** | | | | | |
| Ventilated | 35/112 (28.1) | 35/114 (28.7) | 1.3 | 16.7 | 1.08 [0.57-2.03] |
| On oxygen | 92/759 (11.1) | 95/719 (13.2) | -6.9 | 46.1 | 0.86 [0.59-1.26] |
| Neither | 21/528 (4.0) | 16/539 (2.6) | 3.2 | 9.2 | 1.42 [0.61-3.33] |
| **Bilateral lung lesions** | | | | | |
| Not imaged at entry | 31/179 (17.0) | 24/171 (13.8) | 2.2 | 13.4 | 1.18 [0.58-2.37] |
| Yes | 111/985 (10.2) | 110/945 (11.3) | -0.7 | 54.3 | 0.99 [0.70-1.40] |
| No | 6/235 (2.1) | 12/256 (4.4) | -2.3 | 3.9 | 0.55 [0.15-2.02] |
| **Current smoking** | | | | | |
| Yes | 14/141 (9.2) | 12/124 (9.0) | 0.6 | 5.7 | 1.11 [0.38-3.27] |
| No | 134/1258 (9.8) | 134/1248 (10.5) | -1.1 | 66.1 | 0.98 [0.72-1.35] |
| **Diabetes** | | | | | |
| Yes | 44/341 (12.4) | 33/324 (9.6) | 1.9 | 18.7 | 1.10 [0.61-2.00] |
| No | 104/1058 (8.9) | 113/1048 (10.6) | -2.5 | 52.9 | 0.95 [0.67-1.36] |
| **Heart disease** | | | | | |
| Yes | 44/289 (15.0) | 39/290 (13.2) | 4.0 | 20.3 | 1.22 [0.69-2.16] |
| No | 104/1110 (8.4) | 107/1082 (9.6) | -4.7 | 51.9 | 0.91 [0.64-1.31] |
| **Chronic liver disease** | | | | | |
| Yes | 1/15 (6.7) | 4/23 (18.0) | -1.4 | 1.2 | 0.33 [0.03-3.37] |
| No | 147/1384 (9.8) | 142/1349 (10.2) | 0.9 | 71.1 | 1.01 [0.75-1.37] |
| **Chronic lung disease** | | | | | |
| Yes | 15/95 (13.9) | 12/87 (13.9) | 0.7 | 6.4 | 1.12 [0.41-3.09] |
| No | 133/1304 (9.4) | 134/1285 (10.1) | -1.9 | 65.5 | 0.97 [0.71-1.33] |
| **Asthma** | | | | | |
| Yes | 6/65 (7.7) | 7/56 (12.7) | 0.2 | 1.9 | 1.10 [0.17-7.01] |
| No | 142/1334 (9.8) | 139/1316 (10.2) | -0.1 | 69.2 | 1.00 [0.73-1.36] |
| **Corticosteroids at entry or later (ignoring any biases*)** | | | | | |
| Yes | 69/316 (20.6) | 79/328 (23.3) | 1.6 | 35.0 | 1.05 [0.68-1.62] |
| No | 79/1083 (6.5) | 67/1044 (6.2) | 2.1 | 35.9 | 1.06 [0.69-1.63] |
| **Geographic location** | | | | | |
| Europe or Canada | 47/349 (12.1) | 42/350 (11.3) | 2.9 | 21.8 | 1.14 [0.66-1.98] |
| Latin America | 30/145 (17.2) | 34/148 (23.0) | -0.2 | 15.2 | 0.99 [0.51-1.91] |
| Asia and Africa | 71/905 (7.7) | 70/874 (7.8) | -1.5 | 34.2 | 0.96 [0.62-1.49] |
| **Total** | **148/1399 (9.7)** | **146/1372 (10.3)** | **-0.4** | **72.3** | **1.00 [0.79-1.25]** |

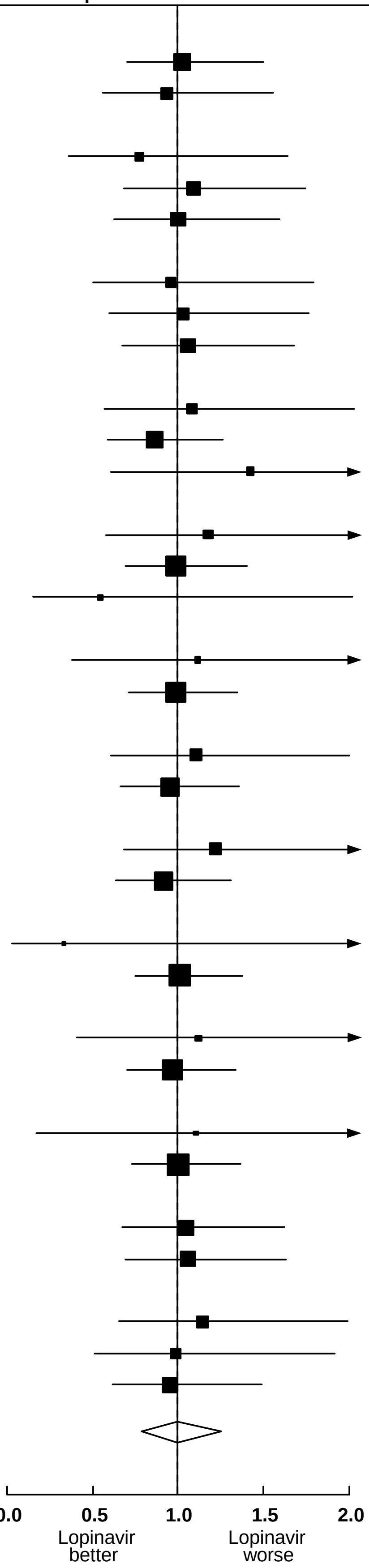


**Heterogeneity test:** $\chi^2_{18}$ **= 8.9; p = 0.96** **2p = 0.97**

99% or 95% confidence interval (CI), K-M Kaplan-Meier.

* RRs in steroid subgroups may be somewhat biased by later steroid use.

#### Figure S9. Rate ratios of any death, stratified by age and respiratory support at entry, Interferon versus Control, by entry characteristics and steroid use at any time

| | Deaths reported / Patients randomized in ITT analyses (28-day risk, K-M%) Interferon | Control | Interferon deaths: log-rank statistics O-E | Variance | Ratio of death rates (RR), & 99% CI (or 95% CI, for total) Interferon : Control |
|---|---|---|---|---|---|
| **Sex** | | | | | |
| Male | 173/1303 (14.4) | 148/1278 (11.9) | 11.8 | 79.3 | 1.16 [0.87-1.55] |
| Female | 70/747 (10.4) | 68/772 (9.6) | 4.5 | 33.5 | 1.14 [0.73-1.78] |
| **Age at entry** | | | | | |
| <50 | 48/720 (7.5) | 35/697 (5.3) | 7.5 | 20.6 | 1.44 [0.82-2.54] |
| 50-69 | 122/934 (14.3) | 108/973 (11.4) | 13.3 | 56.9 | 1.26 [0.90-1.78] |
| 70+ | 73/396 (19.9) | 73/380 (20.9) | -4.0 | 35.8 | 0.89 [0.58-1.38] |
| **Days from hospital admission to randomization** | | | | | |
| 0 | 75/678 (11.6) | 69/677 (10.3) | 5.5 | 34.7 | 1.17 [0.76-1.81] |
| 1 | 70/681 (10.6) | 74/662 (11.0) | -3.4 | 35.5 | 0.91 [0.59-1.40] |
| 2+ | 98/691 (17.0) | 73/711 (11.7) | 14.8 | 41.0 | 1.44 [0.96-2.15] |
| **Respiratory support at entry** | | | | | |
| Ventilated | 55/139 (42.4) | 40/130 (33.8) | 7.7 | 23.0 | 1.40 [0.82-2.40] |
| On oxygen | 178/1429 (14.1) | 163/1430 (11.9) | 10.5 | 84.5 | 1.13 [0.86-1.50] |
| Neither | 10/482 (2.1) | 13/490 (2.9) | -1.3 | 5.7 | 0.80 [0.27-2.35] |
| **Bilateral lung lesions** | | | | | |
| Not imaged at entry | 29/165 (18.7) | 26/177 (15.9) | 3.1 | 13.4 | 1.26 [0.62-2.54] |
| Yes | 206/1723 (13.3) | 184/1718 (11.3) | 13.0 | 96.1 | 1.14 [0.88-1.49] |
| No | 8/162 (4.0) | 6/155 (3.4) | 0.5 | 3.2 | 1.17 [0.28-4.92] |
| **Current smoking** | | | | | |
| Yes | 12/136 (10.3) | 19/138 (14.8) | -2.7 | 7.0 | 0.68 [0.26-1.80] |
| No | 231/1914 (13.1) | 197/1912 (10.7) | 19.5 | 105.7 | 1.20 [0.94-1.55] |
| **Diabetes** | | | | | |
| Yes | 74/489 (16.5) | 58/537 (11.4) | 11.7 | 32.0 | 1.44 [0.92-2.28] |
| No | 169/1561 (11.8) | 158/1513 (10.9) | 3.1 | 80.1 | 1.04 [0.78-1.39] |
| **Heart disease** | | | | | |
| Yes | 58/427 (14.9) | 59/456 (14.2) | 1.3 | 28.3 | 1.05 [0.65-1.70] |
| No | 185/1623 (12.4) | 157/1594 (10.1) | 15.1 | 84.5 | 1.20 [0.90-1.58] |
| **Chronic liver disease** | | | | | |
| Yes | 3/11 (35.2) | 3/22 (15.8) | 0.6 | 0.9 | 1.93 [0.13-29.39] |
| No | 240/2039 (12.8) | 213/2028 (11.0) | 16.1 | 111.8 | 1.15 [0.90-1.47] |
| **Chronic lung disease** | | | | | |
| Yes | 22/114 (20.3) | 22/109 (20.9) | 0.6 | 10.4 | 1.06 [0.48-2.36] |
| No | 221/1936 (12.5) | 194/1941 (10.5) | 15.7 | 102.4 | 1.17 [0.90-1.50] |
| **Asthma** | | | | | |
| Yes | 3/75 (4.5) | 10/97 (10.7) | -1.8 | 2.7 | 0.52 [0.11-2.49] |
| No | 240/1975 (13.3) | 206/1953 (11.0) | 19.4 | 110.1 | 1.19 [0.93-1.52] |
| **Corticosteroids at entry or later (ignoring any biases*)** | | | | | |
| Yes | 166/981 (18.1) | 161/1053 (15.2) | 13.1 | 79.7 | 1.18 [0.88-1.57] |
| No | 77/1069 (7.8) | 55/997 (6.0) | 8.0 | 32.6 | 1.28 [0.81-2.01] |
| **Geographic location** | | | | | |
| Europe or Canada | 27/254 (9.1) | 23/244 (9.0) | 1.7 | 12.3 | 1.15 [0.55-2.40] |
| Latin America | 103/474 (27.1) | 87/478 (18.1) | 14.6 | 45.4 | 1.38 [0.94-2.02] |
| Asia and Africa | 113/1322 (9.3) | 106/1328 (8.8) | 4.1 | 54.1 | 1.08 [0.76-1.53] |
| **Control Type** | | | | | |
| Lopinavir | 87/649 (12.3) | 67/671 (8.9) | 10.5 | 37.9 | 1.32 [0.87-2.01] |
| Standard care | 156/1401 (13.3) | 149/1379 (12.2) | 8.3 | 74.9 | 1.12 [0.83-1.51] |
| **Total** | **243/2050 (12.9)** | **216/2050 (11.0)** | **16.8** | **113.3** | **1.16 [0.96-1.39]** |

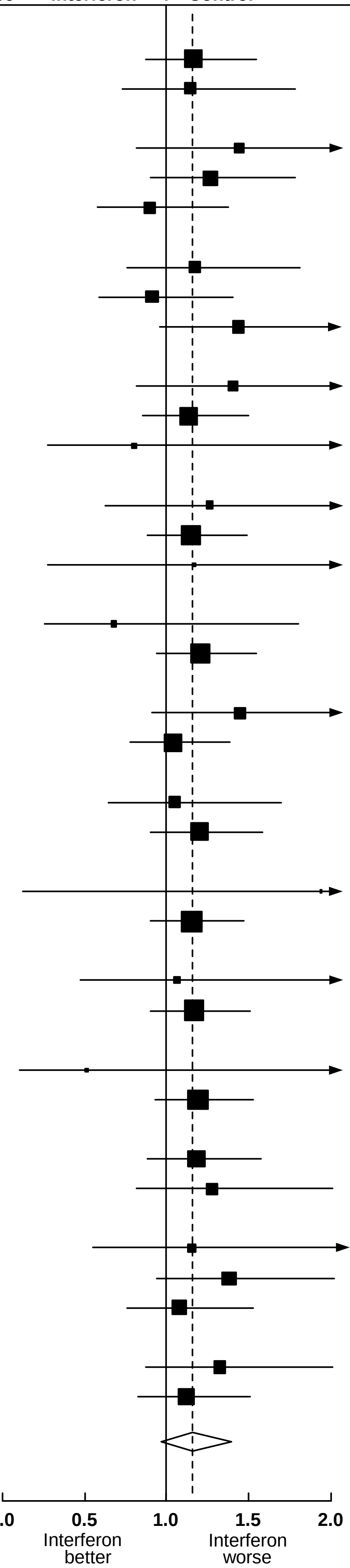


**Heterogeneity test:** $\chi^2_{19}$ **= 19.0; p = 0.46**

**2p = 0.11**

99% or 95% confidence interval (CI), K-M Kaplan-Meier.

* RRs in steroid subgroups may be somewhat biased by later steroid use.

#### Figure S10. RRs for the composite outcome of death or initiation of ventilation: effects of (a) Remdesivir, (b) Hydroxychloroquine, (c) Lopinavir, (d) Interferon, each vs its control

| | Events reported / Patients randomized in ITT analyses (and 28-day risk, %): Active | Control | Active-group events: log-rank statistics: O-E | Variance | Ratio of event rates (RR), & 99% CI (or 95% CI, for total) Active : Control |
|---|---|---|---|---|---|
| **(a) Remdesivir** | | | | | |
| **Age at entry** | | | | | |
| <50 | 114/961 (12.2) | 113/952 (12.4) | 1.0 | 53.2 | 1.02 [0.72-1.45] |
| 50-69 | 240/1282 (20.1) | 245/1287 (20.2) | -6.0 | 111.2 | 0.95 [0.74-1.21] |
| 70+ | 125/500 (27.2) | 121/469 (29.1) | -2.8 | 55.1 | 0.95 [0.67-1.35] |
| **Respiratory support at entry** | | | | | |
| Ventilated | 98/254 (43.0) | 71/233 (37.8) | 7.6 | 40.8 | 1.20 [0.80-1.80] |
| Not ventilated | 381/2489 (16.0) | 408/2475 (17.3) | -15.3 | 178.7 | 0.92 [0.76-1.11] |
| **Total** | **479/2743 (18.5)** | **479/2708 (18.9)** | **-7.7** | **219.5** | **0.97 [0.85-1.10]** |
| Heterogeneity around total $\chi^2_3$: 2.6 | | | | | **2p = 0.60** |
| **(b) Hydroxychloroquine** | | | | | |
| **Age at entry** | | | | | |
| <50 | 29/335 (8.7) | 30/317 (9.6) | -0.2 | 14.1 | 0.99 [0.50-1.96] |
| 50-69 | 79/410 (18.6) | 54/396 (13.2) | 11.1 | 31.2 | 1.43 [0.90-2.27] |
| 70+ | 42/202 (20.4) | 47/193 (24.5) | -3.9 | 20.8 | 0.83 [0.47-1.46] |
| **Respiratory support at entry** | | | | | |
| Ventilated | 35/85 (39.2) | 27/82 (32.3) | 3.4 | 14.8 | 1.26 [0.65-2.46] |
| Not ventilated | 115/862 (13.2) | 104/824 (12.6) | 3.6 | 51.2 | 1.07 [0.75-1.54] |
| **Total** | **150/947 (15.5)** | **131/906 (14.3)** | **7.1** | **66.1** | **1.11 [0.87-1.42]** |
| Heterogeneity around total $\chi^2_3$: 4.3 | | | | | **2p = 0.38** |
| **(c) Lopinavirir** | | | | | |
| **Age at entry** | | | | | |
| <50 | 36/511 (6.9) | 51/501 (9.9) | -7.5 | 21.0 | 0.70 [0.40-1.23] |
| 50-69 | 110/597 (17.8) | 89/596 (14.5) | 9.8 | 45.8 | 1.24 [0.85-1.81] |
| 70+ | 76/291 (25.2) | 81/275 (29.5) | -3.8 | 36.1 | 0.90 [0.59-1.38] |
| **Respiratory support at entry** | | | | | |
| Ventilated | 35/112 (28.1) | 35/114 (28.7) | 1.3 | 16.7 | 1.08 [0.57-2.03] |
| Not ventilated | 187/1287 (14.2) | 186/1258 (14.7) | -2.8 | 86.2 | 0.97 [0.73-1.28] |
| **Total** | **222/1399 (15.3)** | **221/1372 (15.8)** | **-1.5** | **102.9** | **0.99 [0.81-1.20]** |
| Heterogeneity around total $\chi^2_3$: 5.3 | | | | | **2p = 0.88** |
| **(d) Interferon** | | | | | |
| **Age at entry** | | | | | |
| <50 | 68/720 (10.0) | 73/697 (10.6) | -2.4 | 33.5 | 0.93 [0.60-1.45] |
| 50-69 | 177/934 (19.5) | 164/973 (17.4) | 12.3 | 78.3 | 1.17 [0.88-1.57] |
| 70+ | 99/396 (26.1) | 98/380 (26.8) | -3.0 | 44.0 | 0.93 [0.63-1.38] |
| **Respiratory support at entry** | | | | | |
| Ventilated | 55/139 (42.4) | 40/130 (33.8) | 7.7 | 23.0 | 1.40 [0.82-2.40] |
| Not ventilated | 289/1911 (15.7) | 295/1920 (15.6) | -0.8 | 132.8 | 0.99 [0.80-1.24] |
| **Total** | **344/2050 (17.4)** | **335/2050 (16.7)** | **7.0** | **155.8** | **1.05 [0.89-1.22]** |
| Heterogeneity around total $\chi^2_3$: 4.3 | | | | | **2p = 0.58** |

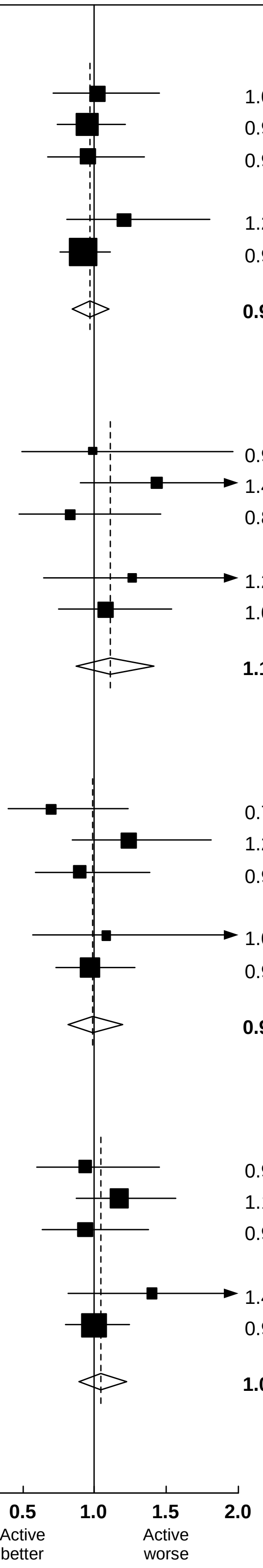


■ 99% or ◇ 95% confidence interval (CI)

**Figure S11. Effects of (a) Remdesivir, (b) Hydroxychloroquine, (c) Lopinavir, (d) Interferon on 14- and 28-day probability of cardiac death**

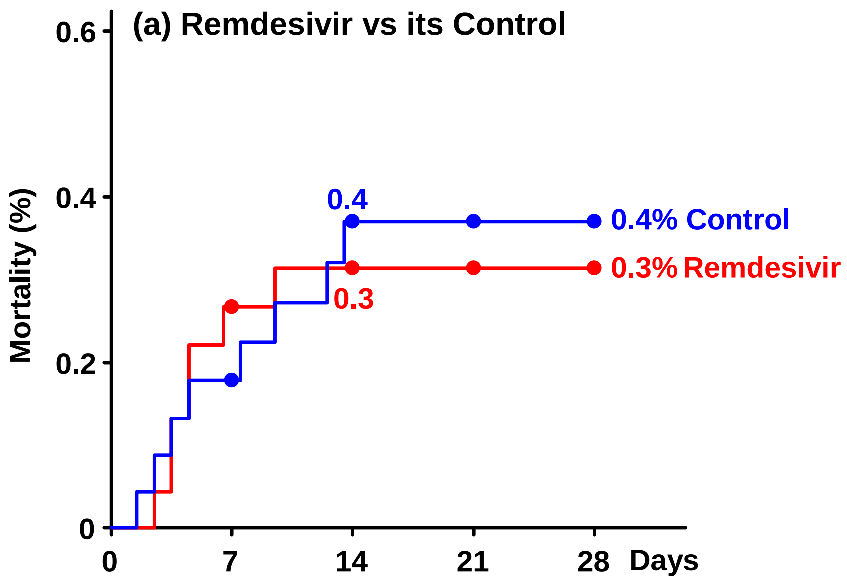


Nos. at risk at start of each week, and nos. dying

| | | | | | | | | | | |
|---|---|---|---|---|---|---|---|---|---|---|
| Remdesivir | 2743 | 6 | 2159 | 1 | 2029 | 0 | 1918 | 0 | 1838 | 0 |
| Control | 2708 | 4 | 2138 | 4 | 2004 | 0 | 1908 | 0 | 1833 | 0 |

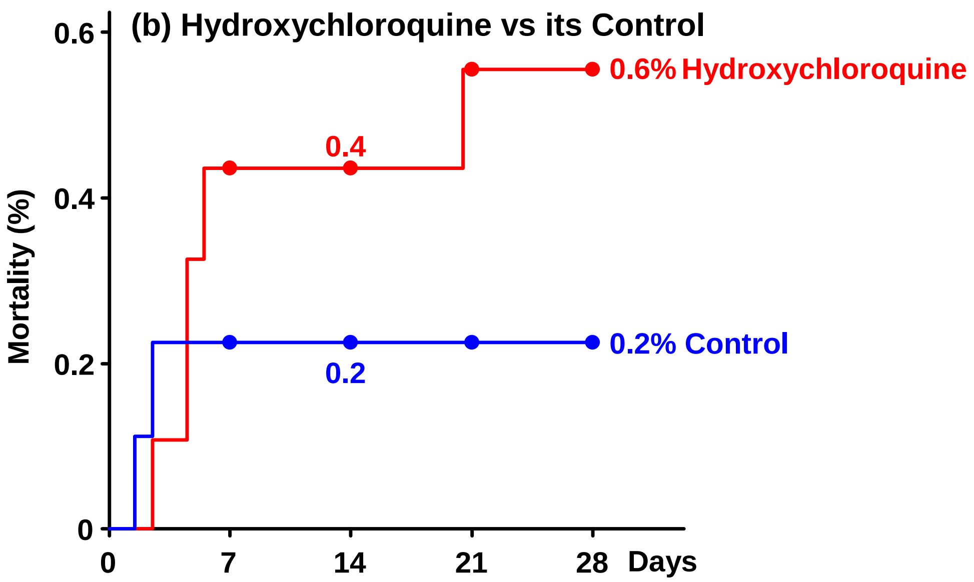


Nos. at risk at start of each week, and nos. dying

| | | | | | | | | | | |
|---|---|---|---|---|---|---|---|---|---|---|
| Hydroxyc . . | 947 | 4 | 889 | 0 | 854 | 1 | 838 | 1 | 833 | 3 |
| Control | 906 | 2 | 853 | 0 | 823 | 0 | 814 | 0 | 809 | 1 |

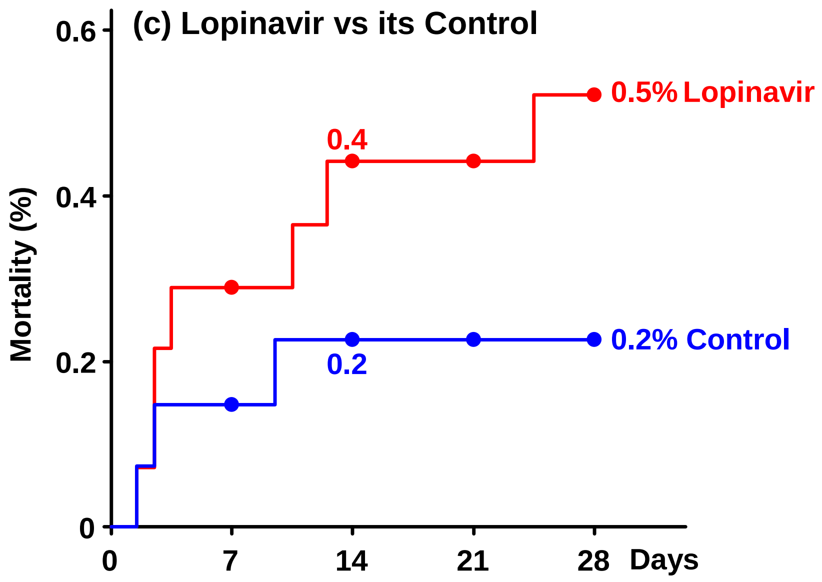


Nos. at risk at start of each week, and nos. dying

| | | | | | | | | | | |
|---|---|---|---|---|---|---|---|---|---|---|
| Lopinavir | 1399 | 4 | 1333 | 2 | 1282 | 0 | 1257 | 1 | 1243 | 0 |
| Control | 1372 | 2 | 1293 | 1 | 1239 | 0 | 1216 | 0 | 1203 | 0 |

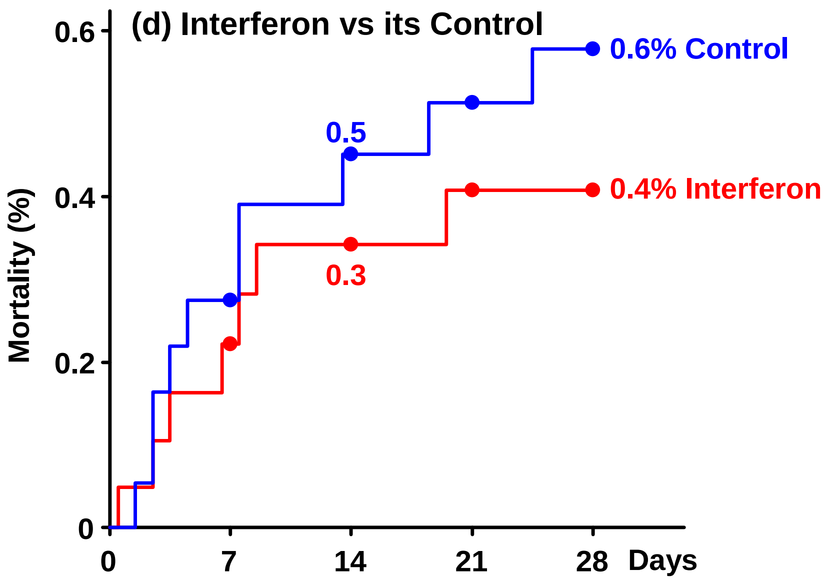


Nos. at risk at start of each week, and nos. dying

| | | | | | | | | | | |
|---|---|---|---|---|---|---|---|---|---|---|
| Interferon | 2050 | 4 | 1669 | 2 | 1554 | 1 | 1483 | 0 | 1410 | 1 |
| Control | 2050 | 5 | 1725 | 3 | 1636 | 1 | 1563 | 1 | 1498 | 0 |
